# Containment measures limit environmental effects on COVID-19 early outbreak dynamics

**DOI:** 10.1101/2020.03.23.20040501

**Authors:** Gentile Francesco Ficetola, Diego Rubolini

**Author notes:** Contributed equally to this work; order was decided with a coin toss.

## Abstract

Environmental factors are well known to affect spatio-temporal patterns of infectious disease outbreaks, but whether the recent rapid spread of COVID-19 across the globe is related to local environmental conditions is highly debated. We assessed the impact of environmental factors (temperature, humidity and air pollution) on the global patterns of COVID-19 early outbreak dynamics during January-May 2020, controlling for several key socio-economic factors and airport connections. We showed that during the earliest phase of the global outbreak (January-March), COVID-19 growth rates were non-linearly related to climate, with fastest spread in regions with a mean temperature of ca. 5°C, and in the most polluted regions. However, environmental effects faded almost completely when considering later outbreaks, in keeping with the progressive enforcement of containment actions. Accordingly, COVID-19 growth rates consistently decreased with stringent containment actions during both early and late outbreaks. Our findings indicate that environmental drivers may have played a role in explaining the early variation among regions in disease spread. With limited policy interventions, seasonal patterns of disease spread might emerge, with temperate regions of both hemispheres being most at risk of severe outbreaks during colder months. Nevertheless, containment measures play a much stronger role and overwhelm impacts of environmental variation, highlighting the key role for policy interventions in curbing COVID-19 diffusion within a given region. If the disease will become seasonal in the next years, information on environmental drivers of COVID-19 can be integrated with epidemiological models to inform forecasting of future outbreak risks and improve management plans.

## 1. Introduction

Host-pathogen interaction dynamics can be significantly affected by environmental conditions, either directly, via e.g. improved pathogen transmission rates, or indirectly, by affecting host susceptibility to pathogen attacks (Altizer et al., 2013). In the case of directly transmitted diseases, such as human influenza and other viral diseases, multiple environmental parameters including local temperatures and humidity impact on virus viability and transmission, with significant consequences for the seasonal and geographic patterns of outbreaks (Shaman and Kohn, 2009; Fuhrmann, 2010; Shaman et al., 2010; Lowen and Steel, 2014; Kampf et al., 2020). The coronavirus SARS-CoV-2 is the aethiological agent of COVID-19, a pandemic zoonosis causing severe pneumonia outbreaks at a global scale (World Health Organization, 2020). During the initial months of 2020, this disease rapidly spread worldwide (Dong et al., 2020), though the early dynamics of COVID-19 outbreaks appeared highly variable. Some countries were experiencing slow growth and spread of COVID-19 cases, while others were suffering widespread community transmission and fast, nearly exponential growth of infections (Dong et al., 2020). Understanding the environmental drivers of early growth rates is pivotal to forecast the potential severity of disease outbreaks and their interactions with containment measures (Britton and Tomba, 2019; Baker et al., 2020; Jung et al., 2020). Given the importance of environmental conditions on the transmission of many pathogens, we tested the hypothesis that the severity of COVID-19 outbreaks across the globe was affected by spatial variation of key environmental factors, and investigated the relative role of environmental conditions and of containment measures adopted by governments on disease spread patterns.

A growing number of studies has been assessing the relationships between COVID-19 growth rate and multiple environmental features, such as temperature, humidity (e.g. Tamerius et al., 2013; Islam et al., 2020a; Kampf et al., 2020; Runkle et al., 2020; Sajadi et al., 2020; Sobral et al., 2020; Wu et al., 2020c), and air pollution (e.g. Bianconi et al., 2020; Rahman et al., 2020; Wu et al., 2020b; Yao et al., 2020; Zhang et al., 2020), while accounting for major socio-economic features of the affected regions (Coelho et al., 2020; Jaffe et al., 2020; Shammi et al., 2020). However, results of these studies were sometimes controversial, casting doubts on the possibility of correctly identifying environmental signals on COVID-19 spread dynamics (Carlson et al., 2020a; Carlson et al., 2020b). Differences among studies can be caused by multiple factors, including lack of standardized methodological framework, differences in spatial extent and scale, and by complex interactions between human transmission, environmental features and containment measures (Baker et al., 2020; Carlson et al., 2020b). Furthermore, both environmental features and containment measures can show complex temporal trends in the course of an outbreak. Studies assessing whether relationships between environment and COVID-19 change are consistent across regions and time periods are pivotal to identify robust and generalizable patterns.

We calculated the mean daily growth rate of confirmed COVID-19 cases during the exponential phase of the epidemic growth curve for the 586 countries/regions (hereafter, regions) (Supplement 1, Fig. S1) where at least 25 cases were reported before June, 2020. Variation at these early epidemic growth rates represents the local progression of the disease and should best reflect the impact of local environmental conditions on disease spread. However, environmental effects on local disease spread could be blurred by containment actions, as in most regions local authorities adopted unprecedented containment measures well in advance or immediately after the detection of an outbreak to mitigate pathogen spread and community transmission (Hellewell et al., 2020; Maier and Brockmann, 2020; Manenti et al., 2020; Thu et al., 2020).

In this study, we first assessed whether COVID-19 growth rate in different regions of the world was affected by major environmental features (temperature, humidity, fine particulate matter; see Methods), controlling for major socio-economic features of the affected regions. Second, we tested whether the stringency of containment measures limited COVID-19 growth rate at the onset of local outbreaks (Maier and Brockmann, 2020). Among the socio-economic factors potentially affecting SARS-CoV-2 transmission dynamics during early outbreaks, we considered human population size, population density, per capita government health expenditure (hereafter, health expenditure) and age structure (see Methods). The importance of a given region in the global air transportation network was expressed as its eigenvector centrality (Coelho et al., 2020) (hereafter, region centrality; see Methods) while containment measures were synthesized into a stringency index (Hale et al., 2020). Finally, to evaluate whether relationships between environment and COVID-19 change were consistent across regions and time periods, we considered regions experiencing outbreaks from January-March 2020 (when outbreaks mostly started before the implementation of strict containment measures) to late May 2020, when lockdown-type containment actions were often adopted even before local outbreaks started. We predicted that late outbreaks, starting under strict containment measures, should be less severe than those starting under no or limited containment, and that environmental effects on COVID-19 growth rate would fade through time, in pace with a progressive increase of the effect of containment actions.

## 2. Materials and methods

### 2.1 COVID-19 dataset

We downloaded time series of confirmed COVID-19 cases (cumulative growth curves) from the Johns Hopkins University Center For Systems Science and Engineering (JHU-CSSE) GitHub repository (https://github.com/CSSEGISandData/COVID-19/) (Dong et al., 2020). JHU-CSSE reports, for each day since January 22, 2020, confirmed COVID-19 cases at the country level or at the level of significant geographical units belonging to the same country, which we broadly defined here as ‘regions’ (e.g. US states, or China and Canada provinces; Supplement 1, Supplementary methods). Data referring to outbreaks occurring on cruise ships were not considered. The cumulative growth curves were carefully checked and obvious reporting errors (a few occurrences of temporary decreases in the cumulative number of cases) were corrected. Our dataset included confirmed COVID-19 cases up to June 15, 2020. From this dataset, we selected data for all those regions in which local outbreaks were detected up to May 31, 2020 (see *Local outbreaks and COVID-19 cases growth rates*).

Overall, we considered data from 159 countries. We considered sub-national level data for the all the countries of the world for which data were easily accessible from the original sources listed in the JHU-CSSE website (for a total of 17 countries; Table S6). Our final dataset included information on 586 regions (Supplement 1, Fig. S2 and Supplementary methods).

### 2.2 Local outbreaks and COVID-19 cases growth rates

To avoid the biases arising because of incomplete spread of the pathogen, our dataset included only those regions experiencing a local COVID-19 outbreak. Therefore, our results are unaffected by patterns occurring in regions where the pathogen showed a limited number of records (e.g. because of distributional disequilibrium, limited connections with other affected areas, or lack of reporting).

The onset of a local COVID-19 outbreak event was defined as the day when at least 25 confirmed cases were reported in a given region. Visual inspection of growth curves showed that, in most cases, below this threshold the reporting of cases was irregular, or growth was extremely slow for prolonged periods. This approach also allowed us to exclude the first cases, often referring to individuals returning from foreign countries and not reflecting local transmission of the pathogen. We then calculated the daily growth rate *r* of confirmed COVID-19 cases for each region after reaching the 25 confirmed cases threshold following the approach proposed by Hall et al. (2014). The method iteratively fits growth curves on successive intervals of a minimum of 5 data points to identify the exponential phase of a cumulative growth curve, and returns the lag phase, and the onset and end of the exponential growth phase. The lag phase, characterized by very slow growth, is followed by the exponential phase (Supplement 1, Fig. S1). Typically, cumulative growth curves of COVID-19 cases begin with exponential growth in the early phases, which begins to decelerate within ca. 10 days of its beginning (e.g. Supplement 1, Fig. S3; see also Maier and Brockmann, 2020). This pattern is similar to what has been documented for earlier phases of other major infectious disease outbreaks (Viboud et al., 2016). We thus restricted the analyses to those regions for which at least 15 days of data after the outbreak onset were available up to June 15, 2020.

Approaches assuming distributional equilibrium can be inappropriate to model the spread of recently emerged infectious diseases (Carlson et al., 2020a). To avoid this issue, we used a dynamic approach, whereby we modelled the dynamics of disease spread within populations (Hall et al., 2014; Carlson et al., 2020a; Coelho et al., 2020). To this end, we computed the mean daily growth rate of confirmed COVID-19 cases during the exponential phase as *r* = [ln(n cases_day end exp. phase_) - ln(n cases_day start exp. phase_)] / (day end exp. phase – day end exp. phase). We also computed the maximum daily growth rate *r*_max_ during the exponential phase according to Hall et al. (2014). Lag and exponential phase duration, and *r*_max_ were computed through the R package growthrates (Hall et al., 2014). Mean and maximum daily growth rates were strongly positively correlated (Pearson’s correlation coefficient, r = 0.95, n = 586 regions), indicating that our growth rate estimates for a given region were highly consistent irrespective of the method used for calculations. By modelling the exponential phase, this approach allowed to focus on local transmission events occurring within the focal region. The average time interval between the first case and the onset of the exponential phase was 19.5 days (SD = 11.1 days), thus cases representing individuals returning from foreign countries likely have a negligible impact on our growth rate estimates.

### 2.3 Environmental variables

We considered two climatic variables that are known to affect the spread of viral diseases: mean air temperature and specific humidity (water vapor pressure), which is a measure of absolute humidity. Previous studies showed that, for coronaviruses and influenza viruses, survival is generally higher at low temperature and low values of absolute humidity (Lowen et al., 2007; Shaman and Kohn, 2009; Tamerius et al., 2013; Lowen and Steel, 2014; Kampf et al., 2020; Yap et al., 2020). For each region, we obtained the mean daily values for temperature (°C) and specific humidity (g/m^3^) from the ERA5 hourly database (Supplement 1, Supplementary methods).

The latency period of the infection, and the lag time between the onset of symptoms, PCR tests and publication of confirmed cases can be highly variable across patients and across areas of the world. For instance, Li et al. (2020a) suggested a mean incubation period of 4-7 days, but also reported cases with shorter incubation, or with incubation > 14 days. Therefore, we measured the potential impact of temperature and humidity in two alternative time windows. First, we considered a broad time period (30 days) occurring before the end of exponential phase. For this 30-days time period, we computed mean climatic conditions (temperature and humidity during 30 days; including the day of the end of the exponential phase and the preceding 29 days; hereafter: 30-days period) (Supplement 1, Fig. S1). This 30-days period aims at covering all the climatic conditions encountered by the broadest range of confirmed cases. Second, we used a narrower time period, focusing on the most frequent time lags between infection and reporting. Following Jüni et al. (2020), we computed mean climatic values assuming an exposure period for infections starting 14 days before the onset of the follow-up period (in our case the start of the exponential phase) and ending 14 days before the end of the follow-up period (in our case the end of the exponential phase) (hereafter: Δ14 days period) (Supplement 1, Fig. S1).

Besides climate, it has been proposed that other environmental parameters may affect variation of COVID-19 outbreak severity. Air pollution, especially fine atmospheric particulate, may enhance the environmental persistence, transmission and effects of coronaviruses (Bianconi et al., 2020; Zhang et al., 2020). We thus calculated the mean annual concentration of PM2.5 for each region (Supplement 1, Supplementary methods).

### 2.4 Socio-economic variables and airport connections

Among socio-economic predictors, we considered mean human population density (Center for International Earth Science Information Network, 2018) (hereafter, population density, expressed in inhabitants/km^2^), total population size (Center for International Earth Science Information Network, 2018), per capita government health expenditure (in US$; average of 2015-2017 values) (Supplement 1, Supplementary methods). Elderly people are more susceptible to develop severe COVID-19 symptoms (Wu et al., 2020a). We thus obtained for each country an estimate of the proportion of the population aged 65 or older (population 65+).

Human mobility is well known to affect pathogen circulation and spatial dynamics (Pybus et al., 2015), and such an effect has been highlighted also for early SARS-Cov-2 spread (Gatto et al., 2020; Kraemer et al., 2020). We thus considered the potential relationships between global airport connections and COVID-19 growth rate. Highly connected regions may experience a higher ‘propagule pressure’ that increase disease diffusion among hosts, ultimately influencing disease growth rates (Coelho et al., 2020). To investigate whether airport connections affected early COVID-19 growth rates, we computed the eigenvector centrality score for each region (region centrality). Highly connected regions have a higher region centrality score (Bonacich, 1987) (Supplement 1, Supplementary methods).

### 2.5 Stringency of containment measures

For each region, we obtained an index of the overall stringency of COVID-19 containment measures adopted by local authorities in the corresponding country at the onset of a local outbreak (hereafter, stringency index). The stringency index was obtained by combining information for each country from two separate data sources (Supplement 1, Supplementary methods). This index simply record the number and strictness of government response measures, hence a higher stringency score does not necessarily imply that a country’s response is more effective than that of other countries with lower scores (Hale et al., 2020). Nevertheless, the stringency index may be helpful to illustrate the timeline of interventions and to assess whether local governments’ policy responses at outbreak onset had any impact on COVID-19 spread within a given region.

### 2.6 Statistical analyses

We relied on linear mixed models (LMMs) to relate variation of COVID-19 growth rate across regions to environmental and socio-economic/management predictors (temperature, humidity, PM2.5, population density, population size, health expenditure, population 65+, region centrality, stringency index). LMMs are an extension of linear models that allow to take into account non-independence of data (Zuur et al., 2009). In our study case, multiple regions within a given country were considered as non-independent as they share multiple features (e.g. health policy, monitoring protocols, economic features other than those considered in the analyses). Country identity was thus included as a random factor to account for non-independence of growth rates from regions belonging to the same country. Non-linear relationships between climatic factors and ecological variables are frequent (Legendre and Legendre, 2012), and have also been suggested for relationships between SARS-CoV-2 occurrence and climate (e.g. Runkle et al., 2020). As in exploratory plots we detected a clear non-linear relationship between *r-*values and climate variables, we included in models both linear and quadratic terms. Humidity, PM2.5, population density, population size, health expenditure and region centrality were log_10_-transformed to reduce skewness and improve normality of residuals. Regression models can be heavily affected by strong collinearity among predictors (|r| ∼0.70 or above) (Dormann et al., 2013). In our dataset, temperature and humidity showed a very strong positive correlation (Supplement 1, Fig. S7 and Table S1). We thus fitted separate models for temperature and humidity, and for different combinations of strongly correlated socio-economic predictors (Supplement 1, Supplementary methods and Table S2).

To assess temporal variation in the importance of different predictors on COVID-19 growth rates, we fitted LMMs considering regions experiencing outbreaks in different periods. Each LMM included data from regions experiencing outbreaks up to a given day. We started from regions experiencing local outbreaks up to February 27, the first day when local outbreaks occurred in at least 50 regions (n = 51 regions), and proceeded on a day-by-day basis until we included all regions experiencing outbreaks up to May 31, 2020 (n = 586 regions; see the cumulative curve in Supplement 1, Fig. S4). The partial *R*^2^ statistic (variance explained by each fixed effect, or semi-partial *R*^2^) was taken as a measure of the importance of each fixed effect in each of these models. Furthermore, we assessed temporal variation of standardized regression coefficients for models fitted at different time points. Airport connections are expected to affect the first phases of the epidemic events, and we therefore tested the effect of region centrality in a model including data up to March 15, 2020 (Supplement 1, Supplementary results). To confirm the time lag period used for the calculation of temperature and humidity (30-days period vs. Δ14 days period) did not affect our results, we repeated analyses twice, first using the 30-days period data, and then using the Δ14 days period data. Climate variables calculated using the 30-days and the Δ14 days periods showed almost perfect correlation across regions (temperature, r = 0.99; humidity, r = 0.99; n = 586 regions).

LMMs were fitted using the lmer function of the *lme4* R package, while tests statistics were calculated using the lmerTest package. Partial *R*^2^ was computed using the *r2glmm* R package. Finally, we used a generalized additive model (GAMs, fitted with the R *mgcv* package) to evaluate the temporal trend of the stringency index at the outbreak date across regions experiencing outbreaks in different periods. For this analysis we used GAMs as we expected a complex temporal pattern and we did not have *a priori* expectations on the shape of relationship between stringency index and time.

## 3. Results

COVID-19 growth rates showed high variability at the global scale (Supplement 1, Fig. S2). The observed daily growth rate during the exponential phase was on average 0.22 (SD = 0.11, N = 586 regions), and ranged from < 0.01 (Argentina, Santiago del Estero and Canada, Prince Edward Island) to 0.72 (Denmark). The exponential growth phase lasted on average 9.0 d (SD = 5.7) and was generally followed by a deceleration of growth, likely as a progressive effect of containment actions and/or increasing awareness by local communities (Supplement 1, Fig. S3) (Maier and Brockmann, 2020). The highest growth rates were observed in temperate regions of the Northern Hemisphere, although relatively fast growth also occurred in some tropical countries, notably Brazil, Indonesia and the Philippines (Supplement 1, Fig. S2). COVID-19 growth rates tended to decrease markedly from March to May (Fig. 1a). At the same time, the stringency of containment measures strongly increased: since the end of March, most outbreaks occurred in regions already under strict containment regimes (Fig. 1b).

**Fig. 1.**
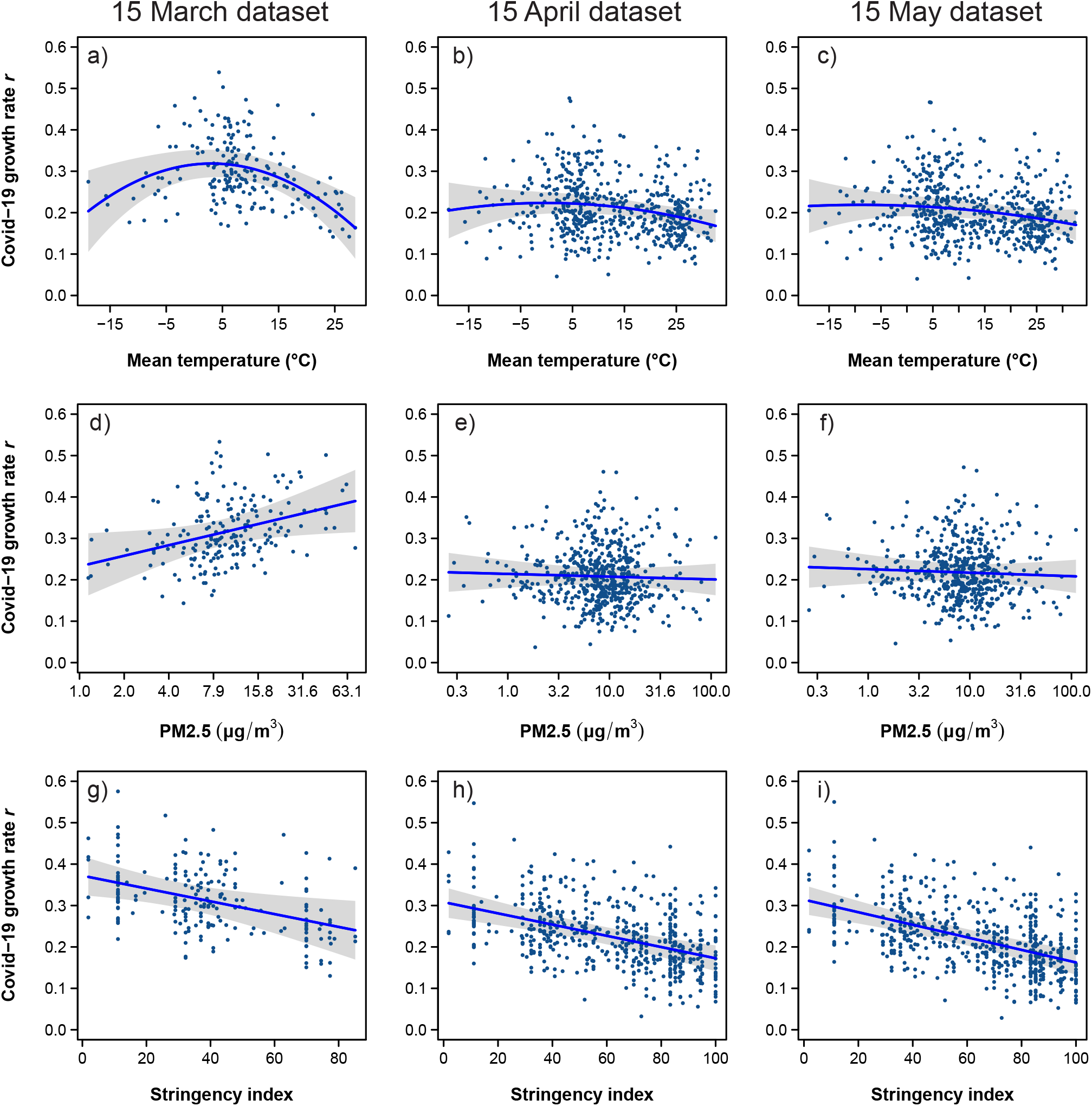
COVID-19 growth rate (a) and stringency of containment measures (b) in regions experiencing COVID-19 outbreaks in different periods. The bold lines represent the fit of a generalized additive model, the shaded area its 95% confidence band. The figures report data for regions where outbreaks occurred between February 27 and May 31, 2020, as before that date data were sparse (< 50 regions experienced outbreaks between January 22 and February 26).

Mixed models including environmental and socio-economic variables explained well variation of COVID-19 growth rate across regions (Supplement 1, Fig. S4). Due to collinearity among predictors (Supplement 1, Table S1), we explored different model formulations (Supplement 1, Table S2 and Fig. S4). The model including temperature (either 30-days period or Δ14 days period), its squared term and PM2.5 as environmental variables, and population density, population size and health expenditure as socio-economic predictors showed the best fit during the early outbreaks, and had similar explanatory power to alternative model formulations when we considered later periods (Supplement 1, Fig. S4). We therefore rely on this model as the main basis for subsequent inference.

Temperature was the strongest environmental predictor during early outbreaks, explaining as much as 20% of the variance in COVID-19 growth rates (Fig. 2). Its effect began to fade when we also included the outbreaks occurring in late March and became negligible from mid-April onward (Fig. 2). PM2.5 exhibited a similar pattern, but its effect size was weaker compared to temperature (Fig. 2). Higher PM2.5 levels were associated with fast growth rates when considering early outbreaks only (Fig. 3). Population size and health expenditure were the strongest socio-economic predictors of growth rates (Fig. 2), the highest growth rates being consistently associated with larger population size and greater health expenditure during both early and late outbreaks (Fig. 3). The stringency of containment measures at outbreak onset consistently negatively predicted COVID-19 growth rates (Fig. 3), becoming the predictor with the strongest effect on growth rates from mid-April onwards (Fig. 2). Results obtained using either the 30-days or the Δ14 days period were nearly identical (Table S3a-b), even though the model using the 30-days period showed slightly higher fit, and temperature effects during early outbreaks were somewhat stronger when considering the 30-days period compared to the Δ14 days period (Fig. 2).

**Fig. 2.**
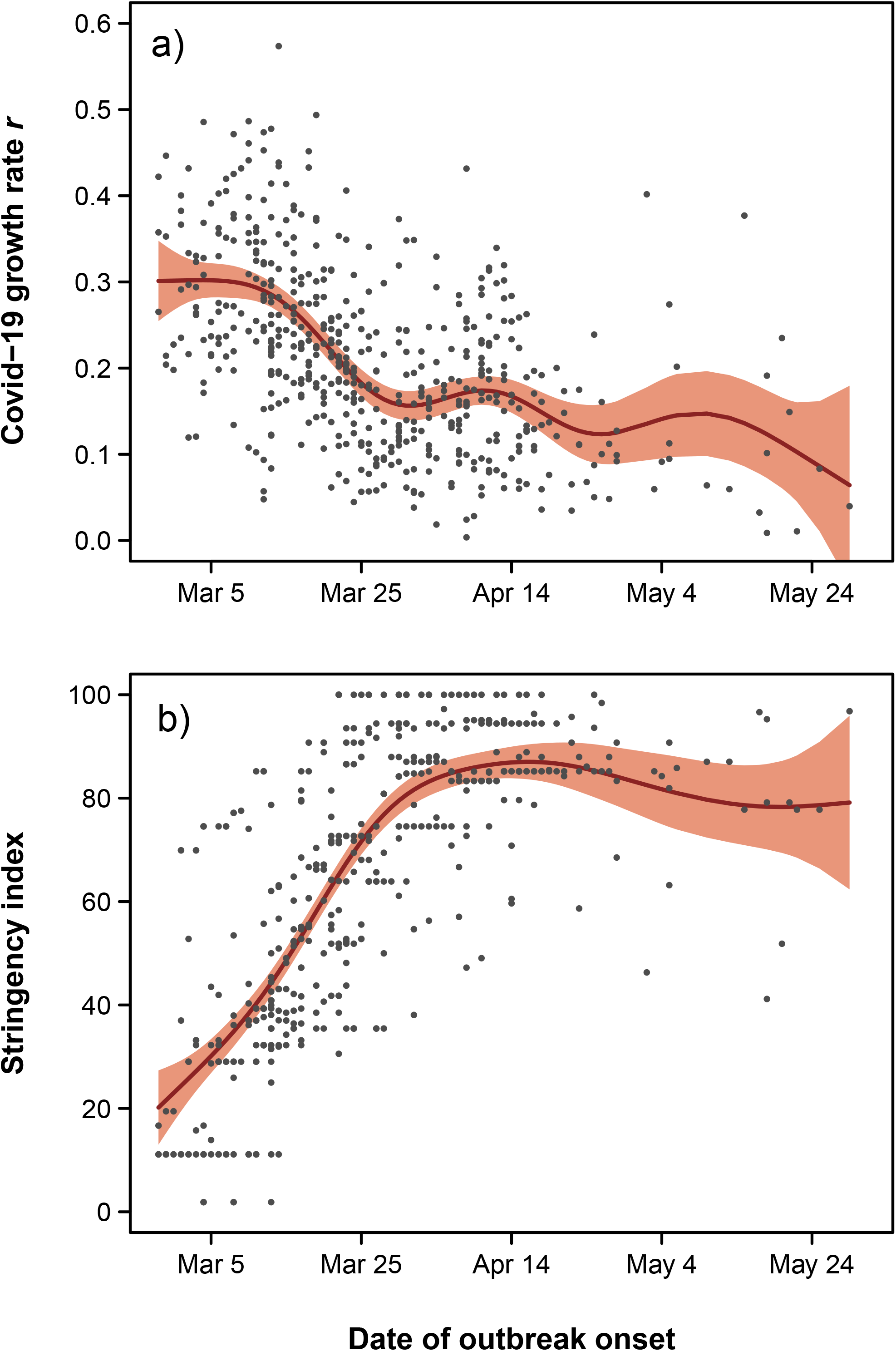
Temporal variation of the importance of variables in explaining COVID-19 growth rate. We fitted regression models starting from regions experiencing outbreaks up to February 27, until we included all regions experiencing outbreaks up to May 31, 2020 (n = 586 regions). The partial *R*^2^ statistic (variance explained by each fixed effect) was taken as a measure of the relative importance of variables. a) temperature calculated using the 30-days period; b) temperature calculated using the Δ14 days period (see Supplement 1, Fig. S1 for details).

**Fig. 3.**
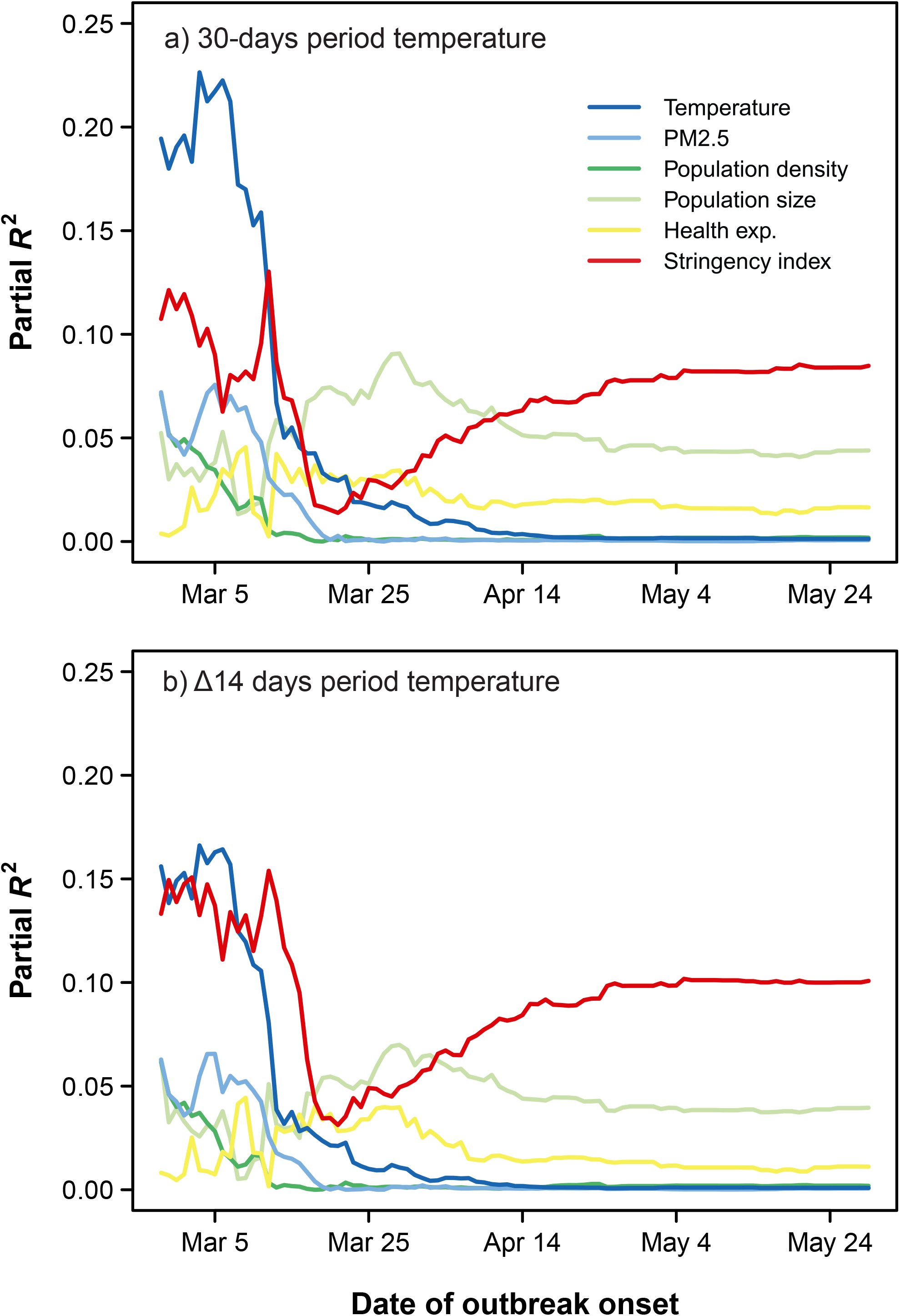
Temporal variation of the relationships between independent variables and COVID-19 growth rate (standardized coefficients). We fitted regression models starting from regions experiencing outbreaks up to February 27, until we included all regions experiencing outbreaks up to May 31, 2020 (n = 586 regions). The plot includes temperature calculated using the 30-days period; the pattern was identical if a Δ14 days period was used (see Supplement 1, Fig. S1 for details). Shaded areas represent 95% confidence bands. When confidence bands do not cross the horizontal broken line (0 threshold), the effect of a given variable is statistically significant (*P* < 0.05).

To illustrate the relationships between COVID-19 growth rate and environmental variables, socio-economic variables, or stringency index, we produced partial regression plots from models fitted on data up to three time points (March 15, to April 15 and May 15; Fig. 4, Supplement 1, Fig. S5; see Supplement 1, Table S3a for model details). For outbreaks occurring up to March 15, growth rates peaked in regions with mean temperature of ca. 5° C, decreasing in both warmer and colder climates (Fig. 4a). Furthermore, highly polluted regions experienced a faster disease spread (Fig. 4d). The effects of temperature and air pollution faded completely when including later outbreaks (Fig. 4c-4f). Higher stringency of containment measures consistently reduced growth rates at all three time points (Fig. 4g-i). Considering the effect of airport connections during early outbreaks or considering alternative environmental and socio-economic variables (absolute humidity, age structure) did not qualitatively alter these conclusions (Supplement 1, Supplementary results and Tables S4-S5).

**Fig. 4.**
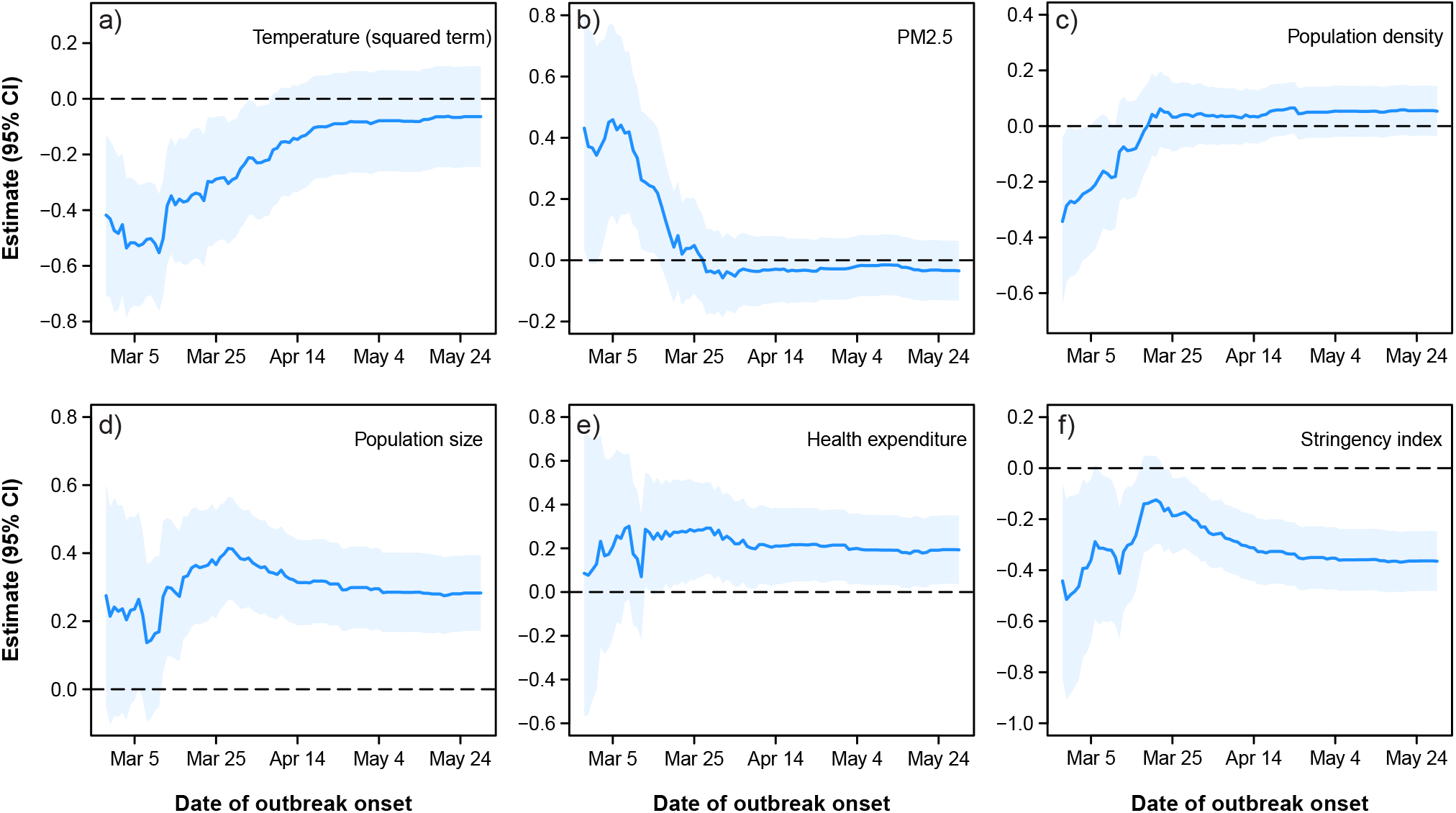
Variation of COVID-19 growth rate in relation to local mean temperature (30-days period), air pollution (PM 2.5) and stringency of containment measures. Partial regression plots from mixed models of COVID-19 mean daily growth rates fitted for local outbreaks occurring up to March 15 (n = 195 regions), April 15 (n = 529 regions) and May 15 (n = 577 regions) are shown. The shaded areas are 95% confidence bands.

## 4. Discussion

The role of environmental drivers on COVID-19 spatial patterns and growth rate is controversial (Araújo et al., 2020; Carlson et al., 2020a; Carlson et al., 2020b; National Academies of Sciences Engineering and Medicine, 2020). Some authors suggested that this disease had a reduced impact and spread in warm climates, and in areas with low pollution and experiencing intense UV radiation (Merow and Urban, 2020; Rahman et al., 2020; Runkle et al., 2020; Sajadi et al., 2020; Sobral et al., 2020; Wu et al., 2020b; Wu et al., 2020c; Zhang et al., 2020), while others reported that socio-economic factors and airport connections have a much stronger impact than environmental drivers (Coelho et al., 2020; Jaffe et al., 2020).

Our results considering the earliest COVID-19 data only (up to March, 2020) are in line with initial evidence reporting less COVID-19 daily new cases and mortality in warm climates (Wu et al., 2020c; Zhang et al., 2020), but exploring a broader time window explained the inconsistency of results across studies. Many previous studies did not explicitly model non-linear effects of climate, and were mostly restricted to the early phase of the global outbreak (Jüni et al., 2020; Wu et al., 2020c). We instead included outbreaks occurring up to the end of May, when COVID-19 reached an almost global spread (Supplement 1, Fig. S2), and adopted an objective approach to identify the exponential phase of outbreaks (Hall et al., 2014). This allowed focusing on early phases of the outbreaks (Maier and Brockmann, 2020), and maximized the possibility of identifying environmental drivers before policy interventions became effective (Merow and Urban, 2020). Finally, we explicitly modeled the spread dynamics within regions (Carlson et al., 2020a; Coelho et al., 2020), thus avoiding the limitations of approaches assuming distributional equilibrium between the pathogen and the environment (Chipperfield et al., 2020).

Multiple non-exclusive processes could explain temperature effects on COVID-19 early growth rate (Araújo et al., 2020; Sajadi et al., 2020). First, the persistence of SARS-Cov-2 and other coronaviruses outside the hosts decreases at high temperature, medium-high humidity, and under sunlight (Lowen et al., 2007; Chin et al., 2020; Kampf et al., 2020; Yap et al., 2020). Second, host susceptibility can be higher in cold and dry environments, for instance because of a slower mucociliary clearance, or a decreased host immune function under harsher conditions (Fares, 2013; Tamerius et al., 2013; Lowen and Steel, 2014). Although SARS-CoV-2 is largely transmitted indoor (Al Huraimel et al., 2020), climatic variation affects host immune response and disease susceptibility (Tamerius et al., 2013). Moreover, it modulates human host behavior, with cold temperatures leading to more time spent indoor and higher disease transmission risk (Tucker and Gilliland, 2007; Fares, 2013; but see also Azuma et al., 2020 for a pattern where contact among people increase in warm days). Thus, climate allows predictions of outbreaks of respiratory illnesses (Shaman et al., 2010; Tamerius et al., 2013), acting both as direct and/or indirect effect. The non-linear relationships between COVID-19 growth rate and temperature detected for early outbreaks (Fig. 4a) might be explained by complex interplays between weather-related changes in human social behavior, changes in host susceptibility to the virus, or changes in virus survival and transmission patterns (Fares, 2013). Overall, with no or weak containment measures, seasonal climatic variation may affect the spatial spread and the risk of severe COVID-19 outbreaks (Merow and Urban, 2020; Wu et al., 2020c), as observed for other viral diseases (Shaman et al., 2010; Tamerius et al., 2013; Lowen and Steel, 2014; Baker et al., 2020), for which seasonal oscillations might lead to the worse outcomes during the colder (autumn-winter) months. Nevertheless, containment measures are able to successfully limit COVID-19 outbreaks in all climatic conditions (Maier and Brockmann, 2020), and climate alone is unlikely to accurately predict transmission in future outbreaks.

The effect of air pollution on COVID-19 spread during early outbreaks was weaker than the effect of local climate. In the early stages of the global outbreak, we observed more severe outbreaks in regions with poor air quality, as gauged by their higher PM2.5 levels, in line with studies suggesting that poor air quality may enhance local transmission and may increase COVID-19 related mortality, possibly not independently of local meteorological conditions (Azuma et al., 2020; Bianconi et al., 2020; Rahman et al., 2020; Wu et al., 2020b; Yao et al., 2020; Zhang et al., 2020). Air pollution can influence COVID-19 spread through different pathways. First, several studies have shown a worsening of respiratory symptoms from viral diseases in populations exposed to poor air quality (Domingo and Rovira, 2020). For instance, chronic exposure to PM 2.5 correlates with overexpression of the alveolar ACE-2 receptor, leading to more severe COVID-19 infection and increasing the likelihood of poor outcomes (Frontera et al., 2020; Wu et al., 2020b). Furthermore, the virus can remain viable in aerosols for some hours, thus high pollution levels might increase its transmission (Frontera et al., 2020). Nevertheless, more studies are required to clarify the actual impact of air pollution on COVID-19 local spread patterns, as well as to identify the actual biological mechanisms (Wu et al., 2020b).

However, the environmental effects on COVID-19 spread during the 2020 global outbreak were not stable through time and disappeared when active containment actions were enforced. Air quality effects became negligible when including outbreaks starting after mid-March, while climate effects lasted a bit longer (until mid-April), but eventually disappeared as well (Fig. 4a-b). From late March onward, most new outbreaks began under severe containment actions (Fig. 1b). A weakening of environmental effects when considering late outbreaks is consistent with the expectation that the enforcement of active containment policies limit the spread potential of the disease and fade associations between climate and disease dynamics (Baker et al., 2020; Maier and Brockmann, 2020).

Analyses of environmental effects on COVID-19 spread have been criticized because SARS-CoV-2 shows a substantial rate of undocumented infections (Li et al., 2020b), and because a high frequency of undocumented cases in some regions (e.g. in Africa) could affect conclusions (Roche et al., 2018; Britton and Tomba, 2019). However, in the early phase of the global outbreak, reported positives largely referred to tested individuals showing COVID-19 symptoms that require hospitalization. Therefore, even though our analyses cannot capture the (unknown) dynamics of asymptomatic infections, they provide information on environmental effects on the spread of symptomatic SARS-CoV-2 cases. Furthermore, our analyses took into account health expenditure, which is strongly correlated to the daily testing rate across countries (Supplement 1, Supplementary methods and Fig. S6). The high COVID-19 growth rate in countries with higher health expenditure likely arose because of more efficient early reporting of cases, thus considering health expenditure in the analyses should at least partly account for differences in testing rate among regions. Finally, we focused on the few days of nearly exponential growth, which generally lasted < 10 days. This limits the possibility that ‘surveillance fatigue’ (Romero-Alvarez et al., 2017) affected our results.

Our analyses provide compelling evidence for the effectiveness of policy interventions in limiting disease spread within regions (Maier and Brockmann, 2020). Although our study was not designed to explicitly test the effect of containment actions, it clearly showed that outbreaks starting under strict containment actions were consistently less severe than those starting under no or weak containment actions. This was already evident for the early (up to end of March) outbreaks, and became the main factor explaining variation in COVID-19 growth rates among countries when considering later outbreaks.

Containing COVID-19 outbreaks is undoubtedly one of the biggest societal challenges. The huge variation of COVID-19 growth rates among regions with similar climate and air quality levels highlights that diverse and complex social and demographic factors, as well as stochasticity, may strongly contribute to the severity of local outbreaks, irrespective of environmental effects. The potential socio-economic drivers of COVID-19 outbreak are many (Coelho et al., 2020; Jaffe et al., 2020). Even if we did not manage to model the spatial spread of the disease across regions, we integrated several variables reflecting potential socio-economic drivers. The positive relationship with human population size might be explained by multiple, non-exclusive processes including an easier control of early outbreaks in regions with small populations, or the occurrence of more trade and people exchanges in the most populated regions, resulting in multiple infection routes and faster spread (Coelho et al., 2020; Jaffe et al., 2020). However, different socio-economic factors were strongly correlated. For instance, areas with high health expenditure were also inhabited by more people older than 65 years (Supplement 1, Table S1), and a linear combination of human population and health expenditure predicts very well international trade of goods and services (Supplement 1, Supplementary methods). Assessing the specific impact of these factors was beyond the aim of this study, but we emphasize that environmental and containment actions effects were consistent irrespective of the specific combination of socio-economic variables being considering, suggesting that unaccounted socio-economic processes should not bias our findings.

In conclusion, our results suggest that local environmental conditions might have affected COVID-19 spread in the early (but not the late) phase of the global outbreak, and that policy interventions can effectively curb disease spread irrespective of environmental conditions (Islam et al., 2020b; Maier and Brockmann, 2020; Thu et al., 2020). Stringent containment measures thus remain pivotal to mitigate the impacts of SARS-Cov-2 infections (Hellewell et al., 2020; Maier and Brockmann, 2020). Yet, information on environmental drivers of COVID-19 can improve the ability of epidemiological models to forecast the risk and time course of future outbreaks, and to suggest adequate preventive or containment actions (Baker et al., 2020). Studies testing the association between environmental features and COVID-19 spread are a rapidly expanding research area that has been attracting increasing attention (Franch-Pardo et al., 2020; Wu et al., 2020b). The unprecedented nature of the pandemic has promoted a growing number of ecological regression analyses, that have identified multiple complex relationships between COVID-19 spread and transmission patterns and diverse environmental features, providing a crucial stimulus to a rapidly evolving area of research (Franch-Pardo et al., 2020; Wu et al., 2020b). The correlative nature of these analyses should call for cautionary interpretations, as identifying the causal processes linking COVID-19 spread dynamics to environmental features remain challenging, still associations detected in ecological analyses can serve as a key starting point for future investigations during the future evolution of the pandemics (Baker et al., 2020; Wu et al., 2020b).

## Supporting information

Complete dataset

## Data Availability

All the relevant data are submitted along with the manuscript

## Acknowledgments

We thank the colleagues of the DISPArati group, especially G. Scarì, for insightful and stimulating discussions. We also thank M. Venegoni, G. Venegoni, V. Longoni, J. G. Cecere and L. Serra for commenting on previous draft of our manuscript, and three anonymous reviewers for insightful suggestions.

## Author contributions

The authors jointly a conceived the work, analyzed data and wrote the manuscript.

## Data and materials availability

All relevant data have been submitted as supplementary files.

