## Supplementary material for "Containment measures limit environmental effects on COVID-19 early outbreak dynamics"

Correspondence:

Francesco Ficetola

**This Supplement 1 includes:**

Supplementary methods

Supplementary results

Figures S1 to S12

Tables S1 to S7

Legends for Datasets S1 to S2

SI References

**Additional supplementary materials for this manuscript include the following:**

Datasets S1 to S2

**Supplementary methods**

**COVID-19 dataset.** Time series of confirmed COVID-19 cases (cumulative growth curves) were downloaded from the Johns Hopkins University Center For Systems Science and Engineering (JHU-CSSE) GitHub repository (https://github.com/CSSEGISandData/COVID-19/). Further details on preliminary data checks are reported in the main text. Data from the Hubei region of China were excluded as the JHU-CSSE dataset does not report cases before January 22, and by that time the epidemic was already largely spreading in Wuhan and nearby municipalities (with 444 confirmed cases for the Hubei region), implying that the early epidemic growth curve was missing. We also excluded data from Nicaragua and Eritrea because of too irregular reporting of confirmed cases. Analyses including these regions yielded results that were highly consistent with those presented in the main text.

We considered sub-national level data for the eight largest countries of the world (Russian Federation, Canada, USA, China, Brazil, Australia, India, Argentina) and for several smaller countries for which data were easily accessible from the original sources listed in the JHU-CSSE website (see Table S6). Furthermore, for Quebec (Canada), we limited our analysis to Southern Quebec data (i.e. health regions south of 49°N), since in the northernmost Quebec health regions (extending to sub-polar areas up to 62° N) nearly no COVID-19 cases were detected before March 21 (see Table S6 for data sources). Sub-national boundaries followed the global database of administrative areas ([www.gadm.org](http://www.gadm.org)). Some small special administrative territories enclosed within larger regions were merged with those regions (Argentina: C.A. Buenos Aires merged with Buenos Aires province; Colombia: Bogotà D.C. merged with Meta region), and the Ladakh region of India was merged with the Jammu and Kashmir Union Territory. Finally, to reduce heterogeneity among regions, we excluded data from those regions with less than 100,000 inhabitants, or with total surface < 1,000 km^2^ and less than 1 million inhabitants. The complete list of regions included in the dataset is reported in Table S7.

**Environmental variables**. We downloaded hourly values of temperature and 2-m dewpoint temperature at the 0.25° spatial resolution from the ERA5 hourly database (https://doi.org/10.24381/cds.adbb2d47); we then calculated specific humidity using the ‘humidity’ package in R (Cai, 2019). Concerning air pollution, we extracted values of annual concentration (µg/m^3^) of ground-level fine particulate matter (PM2.5) for 2016 from the NASA Socioeconomic Data and Applications Center (spatial resolution 0.01°) (van Donkelaar et al., 2018). We performed all spatial analyses using the *raster* package in R (Hijmans, 2019).

**Socioeconomic variables and airport connections**. Human population density (Center for International Earth Science Information Network, 2018) (inhabitants/km^2^), total population size (Center for International Earth Science Information Network, 2018) and per capita government health expenditure were downloaded from the World Health Organization database at https://apps.who.int/nha/database. Data for Kosovo, Montenegro, Libya, Syria, West Bank and Gaza were obtained from http://documents.worldbank.org. Health expenditure was available at country-level only, hence regions within countries were assigned the same health expenditure value. Testing rate is a further factor that can affect the number of detected cases and growth rate, and can covary with health expenditure. We obtained information on daily testing rate (mean value of the daily number of tests/1000 persons performed up to May 31, 2020) from Roser et al. (2020). Information on daily testing rate was available for a subset of 84 countries (Supplementary Data).

Estimates of population 65+ were largely based on the most recent available data retrieved from the United Nations website (https://population.un.org/wpp/Download/Standard/Population/, dataset ‘Population by Age Groups - Both Sexes’). International trade is an additional process that has been proposed to affect COVID-19 dynamics (Jaffe et al., 2020). Following Jaffe et al. (2020), we downloaded data on the total imports of goods and services (for 2017, expressed in US$) from the World Bank Database (https://data.worldbank.org/indicator). However, total import of goods and services was strongly related to a linear combination of total population size and health expenditure (linear model on log-transformed values: *R*^2^ = 86). Furthermore, per-capita import of goods and services was strongly related to health expenditure (Pearson’s correlation, r = 0.88). The strong correlation among these variables prevents their simultaneous inclusion in regression models (Dormann et al., 2013), and makes it difficult to identify the role of individual predictors.

To investigate the importance of airport connections on COVID-19 growth rate, we first constructed an air transportation network based on the OpenFlight database (https://openflights.org/data.html), which reports data for 67,432 direct airline connections among world airports (database updated up to 2014). The location of world airports was obtained from the OpenFlight website and from https://datahub.io/core/airport-codes. Airports were assigned to regions included in the analyses. We then obtained a directed weighted graph with regions as nodes, flight connections among regions as vertices, and number of pairwise connections among regions as weights (Fig. S8). For regions considered up to March 21, 2020, the network included 50,547 direct airline connections among 1,951 airports (Fig. S8). As an index of connectedness of regions, we computed the eigenvector centrality score (region centrality), which is a measure of the influence of a given node in a network (Bonacich, 1987). It estimates the importance of a given region in the network taking into account the number of connections with other regions, whereby connections with highly connected regions contribute more to the region centrality score than connections with low-scoring regions (Bonacich, 1987). Network analyses were performed with the *igraph* package in R (Csardi and Nepusz, 2006).

**Stringency of containment measures**. To assess the stringency of containment measures, we relied on two data sources: 1) the Oxford COVID-19 Government Response Stringency Index (heareafter, Oxford SI), a composite measure released by the University of Oxford (Hale et al., 2020) that is based on nine response indicators including school closures, workplace closures, and travel bans, rescaled to a value from 0 to 100 (100 = strictest response) (https://github.com/OxCGRT/covid-policy-tracker/raw/master/data/OxCGRT_latest.csv; data downloaded on June 25, 2020); and 2) the Assessment Capacities Project (ACAPS) COVID-19 Government Measures Dataset (https://www.acaps.org/covid19-government-measures-dataset), downloaded from the *tidycovid19* R package (Gassen, 2020a), which reports the number of social distancing, of movement restrictions, and of lockdown measures on a daily basis, net of lifted restrictions. The latter three were combined and made comparable to the Oxford SI by appropriately ranking them on a 0-100 scale according to Gassen (2020b) (hereafter, ACAPS SI). Up to May 31, 2020, the correlation between these two stringency indexes was strong (r = 0.78; n = 577 regions for which both indexes were available) (see also (Gassen, 2020b)), even though the Oxford SI yielded somewhat larger stringency values than the ACAPS SI (mean ± SD, 60.5 ± 28.5 vs. 50.7 ± 25.0). Since the Oxford SI was not available for nine countries, we estimated the stringency index at each local outbreak date as the maximum value between the Oxford SI and the ACAPS SI.

**Statistical analyses**. Regression models can be heavily affected by strong collinearity among predictors (|r| ~0.70 or above) (Dormann et al., 2013). In our dataset, temperature and humidity showed a very strong positive correlation (Fig. S7 and Table S1); furthermore, health expenditure was strongly positively related to population 65+, and population size was strongly positively associated with region centrality (Table S1). The remaining predictors showed weaker correlations (Table S1). Therefore, temperature and humidity, and health expenditure and population 65+ could not be considered together in the same models (Dormann et al., 2013; Giam and Olden, 2016). We thus repeated analyses with different combinations of predictors (see summary in Table S2). First, we considered temperature, PM2.5, population size, population density and health expenditure as predictors. Then, we fitted models including population 65+ instead of health expenditure. Finally, we repeated analyses including humidity instead of temperature. All models were fitted using climate variables computed during both a 30-days period and a Δ14 days period (see Fig. S1, Materials and methods and Results in the main text). The effect of region centrality was included in the early model (data up to March 15) instead of population size. The datasets used for the analyses are provided in Dataset S1.

To confirm that spatial autocorrelation did not bias the outcome of our analyses, we calculated the spatial autocorrelation (Moran’s *I*) of the residuals of models including data up to March 15, April 15, and May 15 using the *EcoGenetics* R package (Roser et al., 2017) using a lag distance of 1000 km. The procedure was repeated for all 5 model formulations reported in Table S2. Model residuals did not show significant spatial autocorrelation (in all cases, Moran’s *I* < 0.01, *P* > 0.1), suggesting that spatial autocorrelation was not a major issue in our analyses (Beale et al., 2010).

**Supplementary results**

**Analyses considering humidity instead of temperature as a climatic predictor**. Due to the strong correlation between temperature and humidity (Fig. S7, Table S1), we repeated the analyses reported in the main text including humidity instead of temperature as a climatic predictor (30-days period) of COVID-19 growth rate. Including humidity instead of temperature did not significantly alter our conclusions: humidity and PM2.5 significantly predicted COVID-19 growth rate in the early phase of the global outbreak, whereas their effects faded from mid-April onward (Fig. S9a). The effects of socio-economic variables and stringency index were qualitatively similar to those detailed in the main text (Fig. S9). Details results of parameter estimates and model fits for representative dates (March 15, April 15, May 15) are reported in Table S4. The results were very similar to the models including temperature as a climatic predictor (Tables S3-S4), even though models including temperature provided a better fit to the data compared to those including humidity, especially in the early phase of the global outbreak, as judged by lower AIC values (see Tables S3-S4).

**Analyses considering the proportion of population aged 65 or older**. To account for the potentially higher susceptibility of a relatively old population on COVID-19 growth rates, we repeated the analyses described in the main text (30-days period temperature as the climatic predictor) and in the above paragraph (humidity as the climatic predictor) including in LMMs the proportion of population aged 65 or older (population 65+) instead of health expenditure. These two variables were strongly positively correlated (Table S1) and were thus not included together in the same models. Overall, these models had a slightly poorer fit than those including health expenditure (Fig. S4). The results concerning environmental effects (temperature and PM2.5) were unaltered compared to those presented in the main text or in the above paragraph, and the same was true for the other socio-economic predictors included in the models (Fig. S10-S11). The effect of population 65+ was generally positive, with regions characterized by an older population experiencing somewhat higher growth rates, but the effect was only statistically significant from the end of March onwards (Fig. S10e-S11e). Hence, population 65+ was related to significantly higher local COVID-19 growth rates in the late but not in the early phases of the global outbreak.

**Analyses considering airport connections among regions.** To account for the potential impact of human mobility on COVID-19 growth rates, we refitted the LMM of mean growth rate up to March 15 (with 30-days temperature) including region centrality in the air transportation network instead of population size. These two variables were strongly positively correlated (Table S1) and could thus not be included together in the same models. The model was fitted on data from 186 regions (instead of 195) from 69 countries because 9 regions had no airports or connections (Supplementary methods and Fig. S8). The model confirmed a significant quadratic effect of mean temperature on growth rates, a significant effect of PM2.5 and of the stringency index, whereas region centrality in the global transportation network had a negligible effect (Table S5). Furthermore, this model showed considerably lower support than the one including population size instead of region centrality (ΔAIC = 9.19, based on the same sample of 186 regions), suggesting that effects of region centrality reported in previous studies (Coelho et al., 2020) could be a side-effect of the better connection of regions with large population size.

**Analyses using maximum growth rate instead of mean daily growth rate***.* We repeated the analyses described in the main text (with 30-days period temperature) including in LMMs the maximum daily growth rate during the exponential phase (*r*_max_) instead of the mean daily growth rate *r* (see Fig. S1). Maximum growth rates were on average slightly larger than mean growth rates (*r* = 0.28 ± 0.11 SD, *r*_max_ = 0.30 ± 0.11; see Table S7). The results were nearly identical to those presented in the main text (Fig. S12). The fit of these models was slightly poorer compared to the models including mean growth rate in the early phase of the outbreak, but slightly better during the middle and late phases of the outbreak (blue line in Fig. S4). All main conclusions concerning the effect of different environmental or socio-economic predictors on COVID-19 growth rates remained thus unaltered when considering maximum growth rate instead of mean daily growth rate.

**Fig. S1.** Example of a confirmed COVID-19 cases cumulative growth curve (Thailand; data are shown only up to March 21 for clarity of representation). The first two confirmed cases were reported on January 22, 2020 (day 22). The 25^th^ case threshold was reached on day 35 (February 4). This is the time when the lag phase, with very slow growth, begins (blue dots). The lag phase is followed by an exponential growth of confirmed cases (exponential phase, orange and red dots). After the exponential phase, which here starts at day 69, the growth rate often declined (Fig. S3), possibly because of active containment actions. The orange and red dots identify the period over which the mean daily growth rate *r* of COVID-19 cases was computed; the red dots identify the period (5 days minimum) where the maximum growth rate (*r*_max_) was achieved. We assumed that the growth rates *r* and *r*_max_ achieved during the exponential phase represent proxies of COVID-19 growth in a completely susceptible host population. The periods over which the mean climatic conditions of each country/region were computed are shown: a) 30-days period (29 days before the end of the exponential phase, plus the day of the end of the exponential phase; in this case from day 55 to day 84); b) Δ14 days period (starting 14 days before the onset of the exponential phase and ending 14 days before the end of the exponential phase; in this case from day 55 to day 70; Jüni et al., 2020). Length of phases and growth rates were computed following Hall et al. (2014)

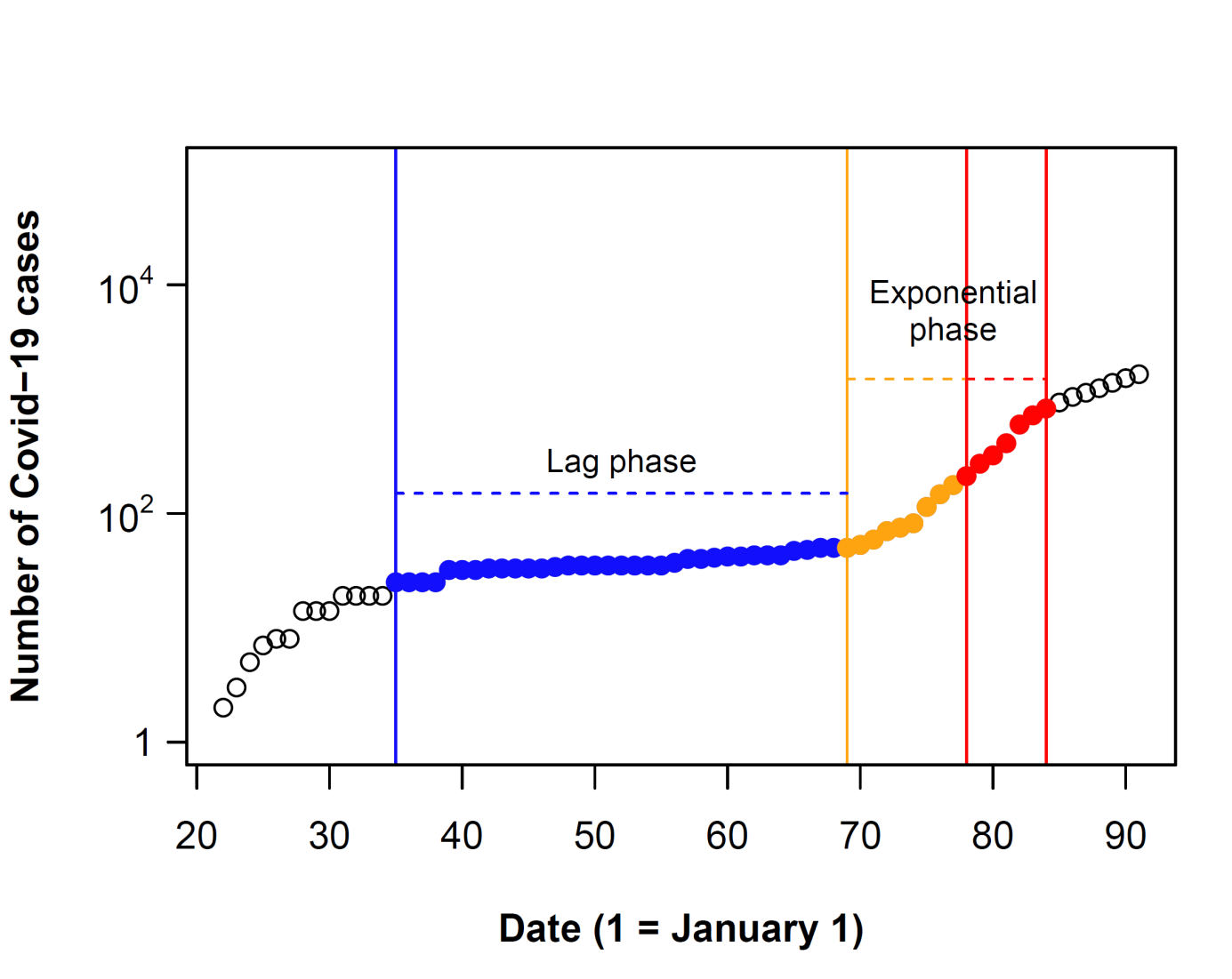

Day 55-84 (30-days period)

**Time periods used to calculate mean climatic conditions:**

Day 55-70 (Δ14 days period)

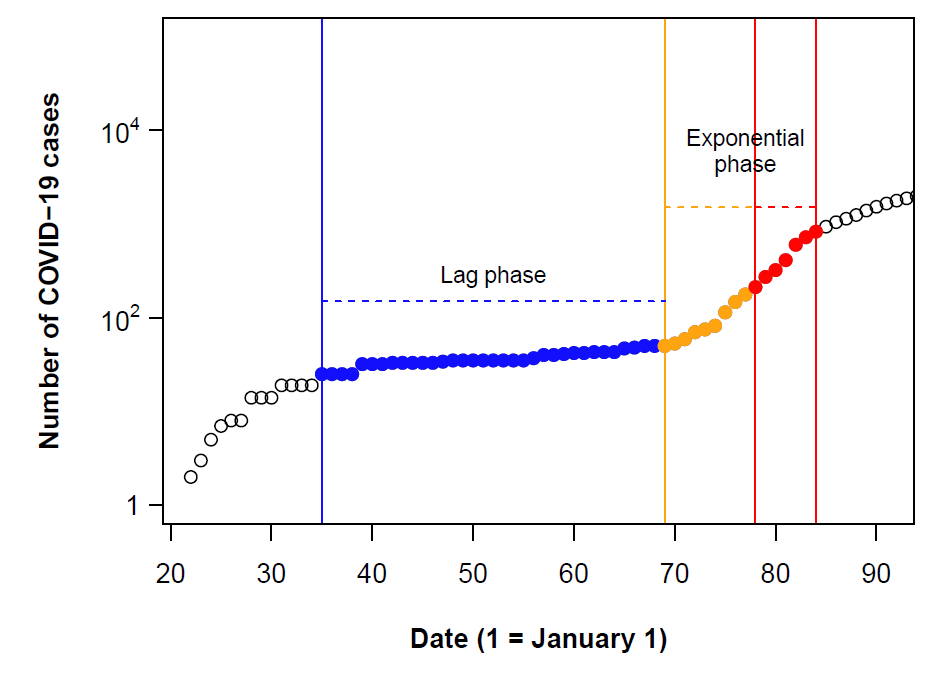

**Fig. S2.** a) Growth rate of confirmed COVID-19 cases in regions where local outbreaks occurred up to May 31, 2020 (n = 586 regions). b) Spatial variation in the date of outbreak onset in the 586 regions.

a)

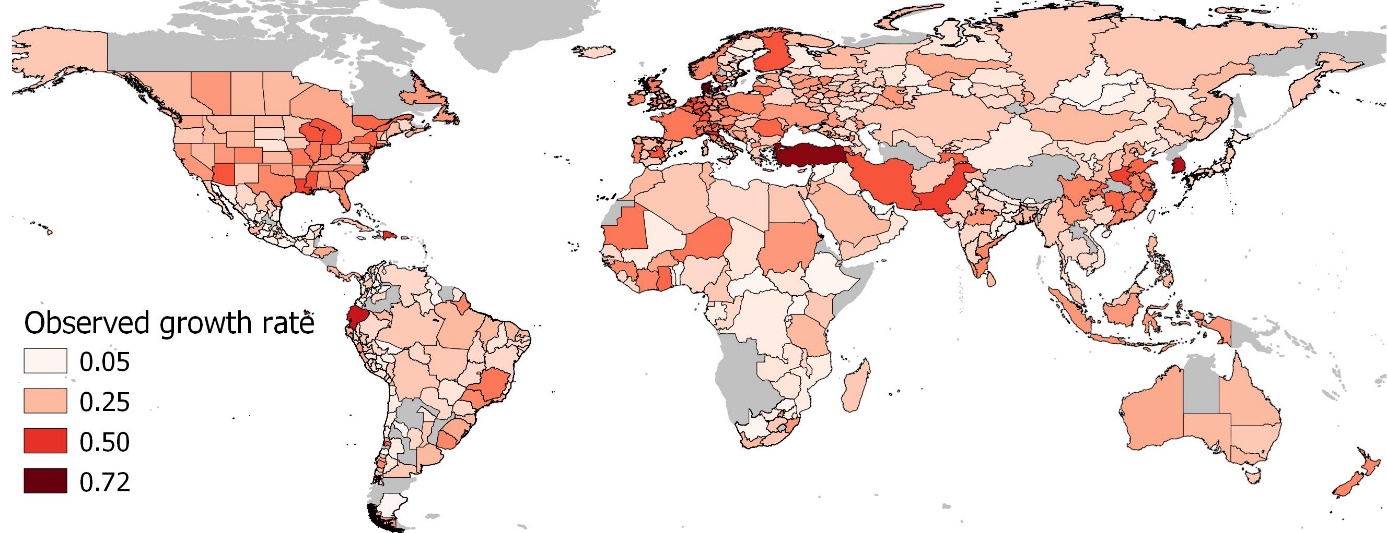

b)

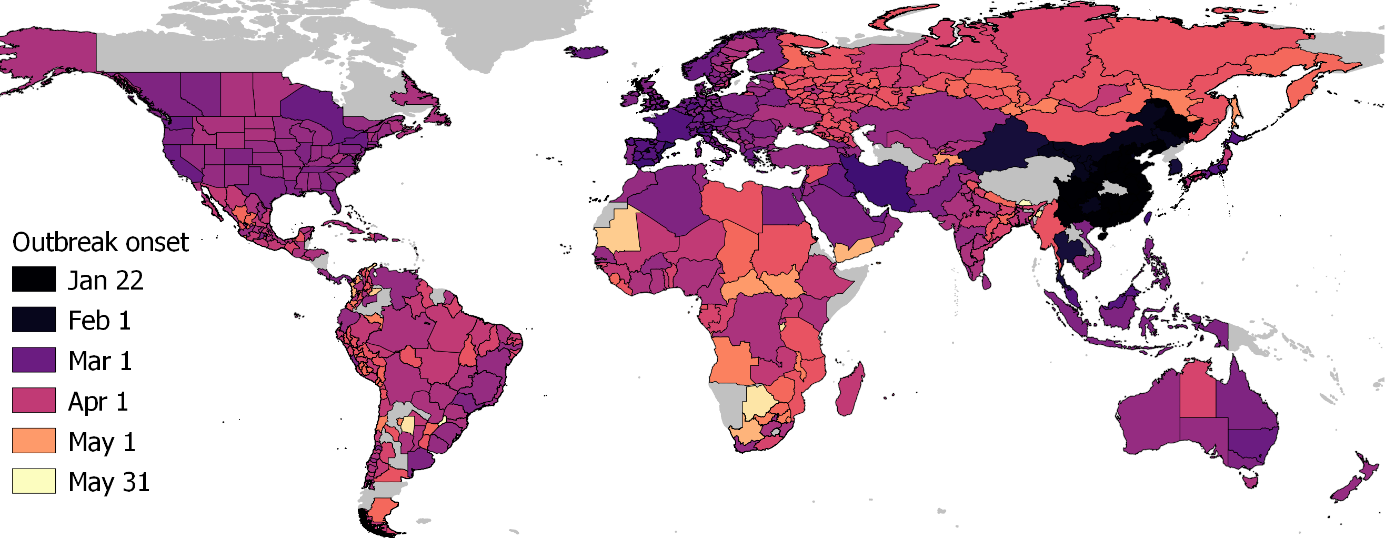

**Fig. S3.** Growth curves up to June 15, 2020 for 20 representative regions. Blue dots: lag phase; orange and red dots: exponential phase; red dots: maximum growth period within the exponential phase.

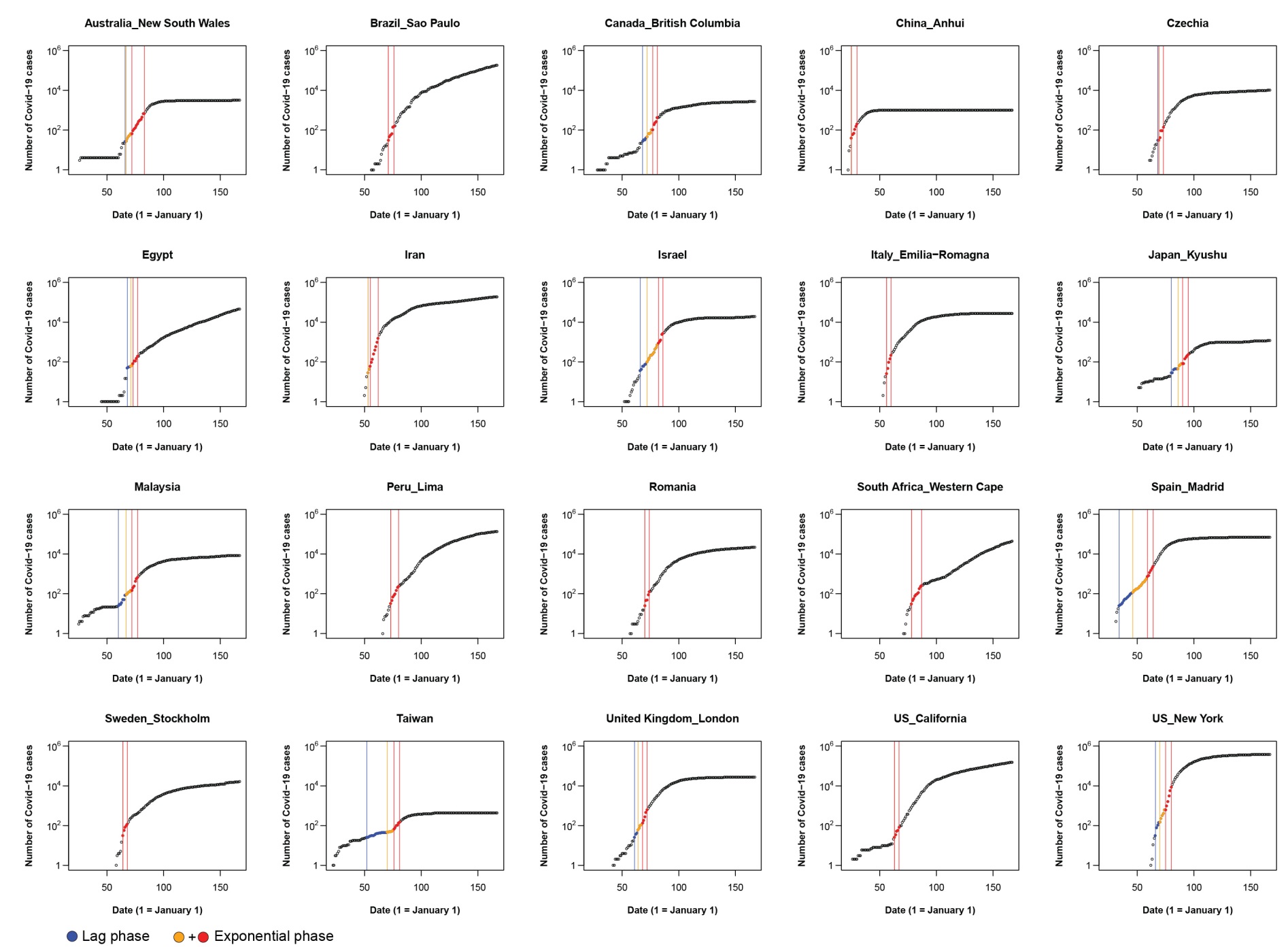

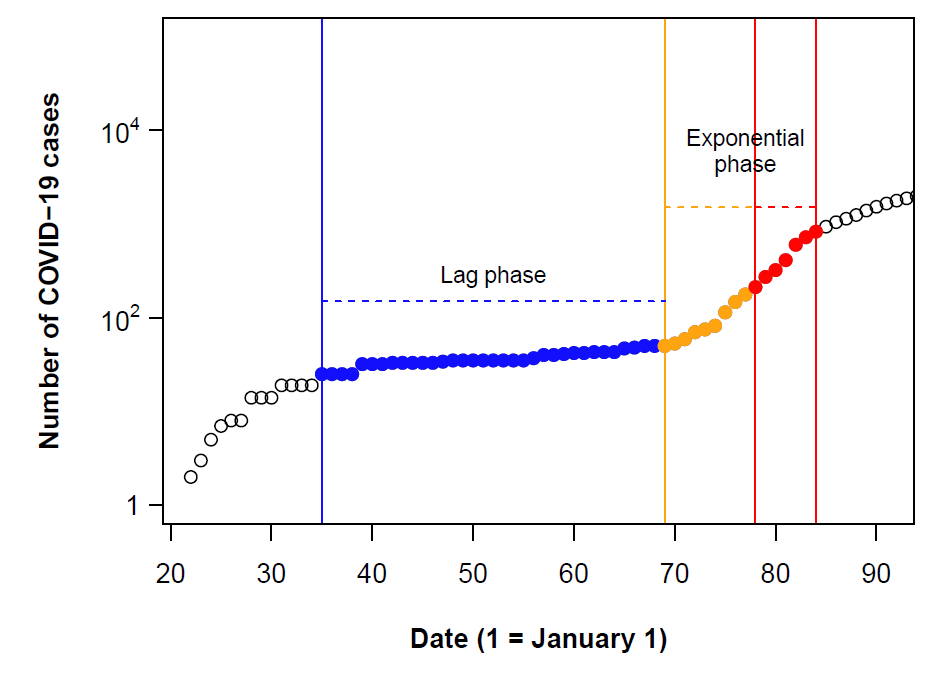

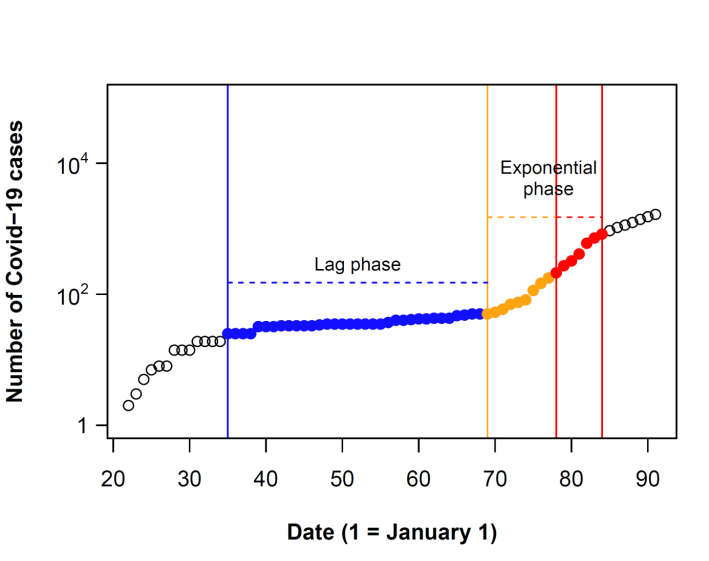

**Fig. S4.** Temporal performance of alternative formulations of models relating COVID-19 growth rate to different combinations of environmental and socio-economic variables. For a given date, the model with highest marginal *R*^2^ shows the best performance in explaining variation of COVID-19 growth rates for outbreaks starting before that date. At a given date, all models include the same number of predictors. Therefore, alternative measures of performance (e.g. AIC) show an identical pattern, with models having the highest marginal *R*^2^ showing the lowest AIC values (see e.g. Tables S3-S4 for AIC values of representative models including temperature or humidity as climatic predictors). The dotted line shows the temporal variation in the number of regions included in the models. See Table S2 for details on the variables included in each model. Upper panel: climatic variables computed over a 30-days period; lower panel: climatic variables computed over a Δ14 days period (see Fig. S1).

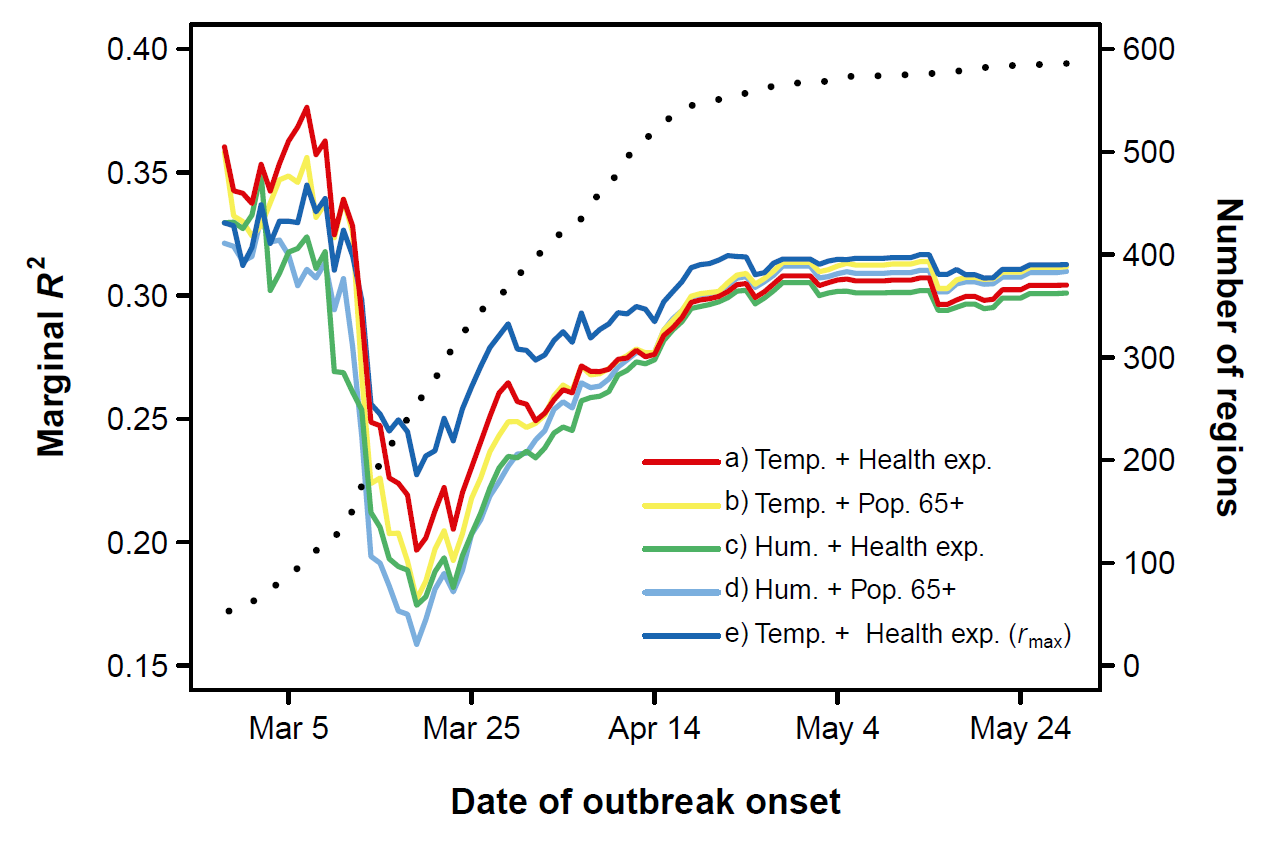

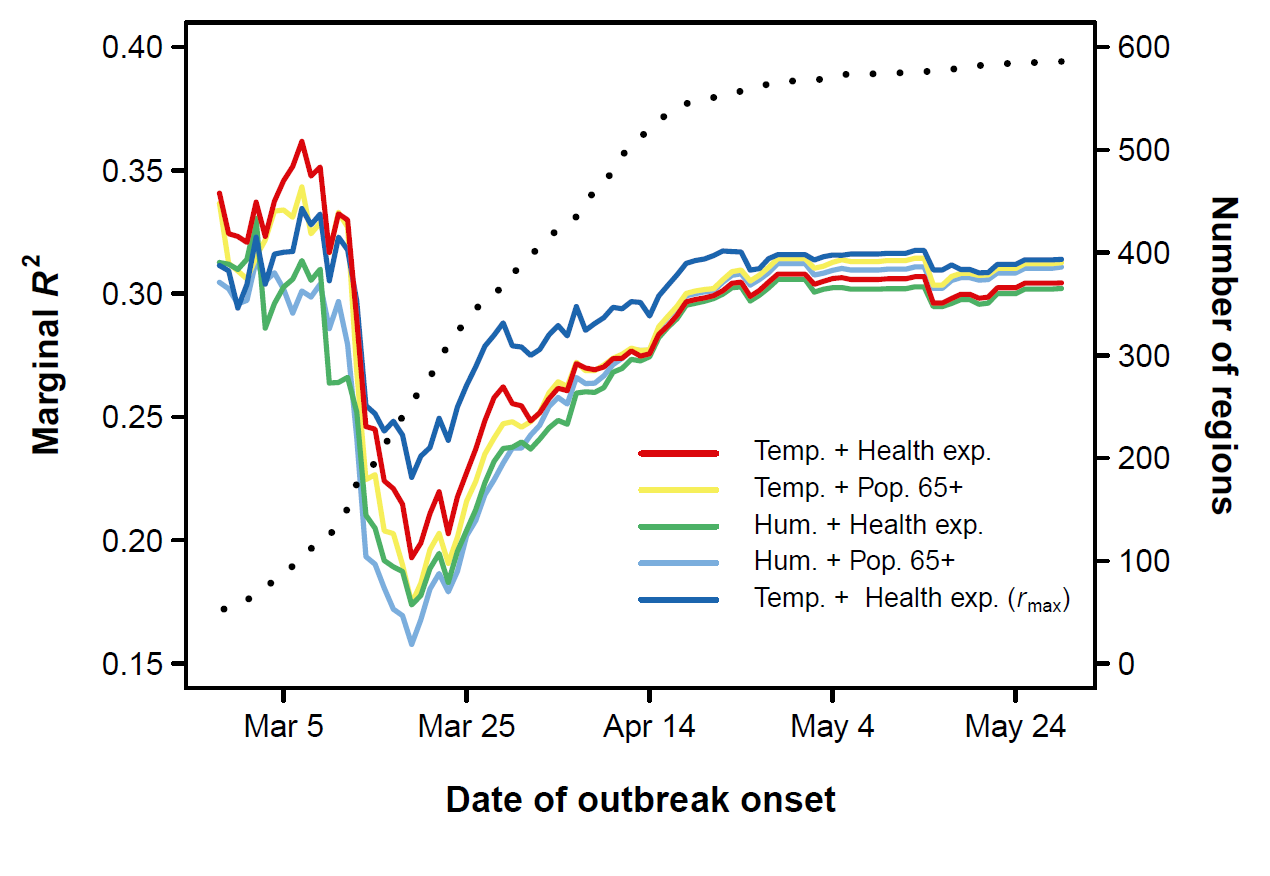

**Fig. S5.** Partial regression plots (Breheney and Burchett, 2017) of the effects of population density, population size and per capita health expenditure (lower panel) from representative mixed models of COVID-19 growth rate including temperature (30-days period) as a climatic predictor (see Table S3a for details of model fits). Shaded areas are 95% confidence bands.

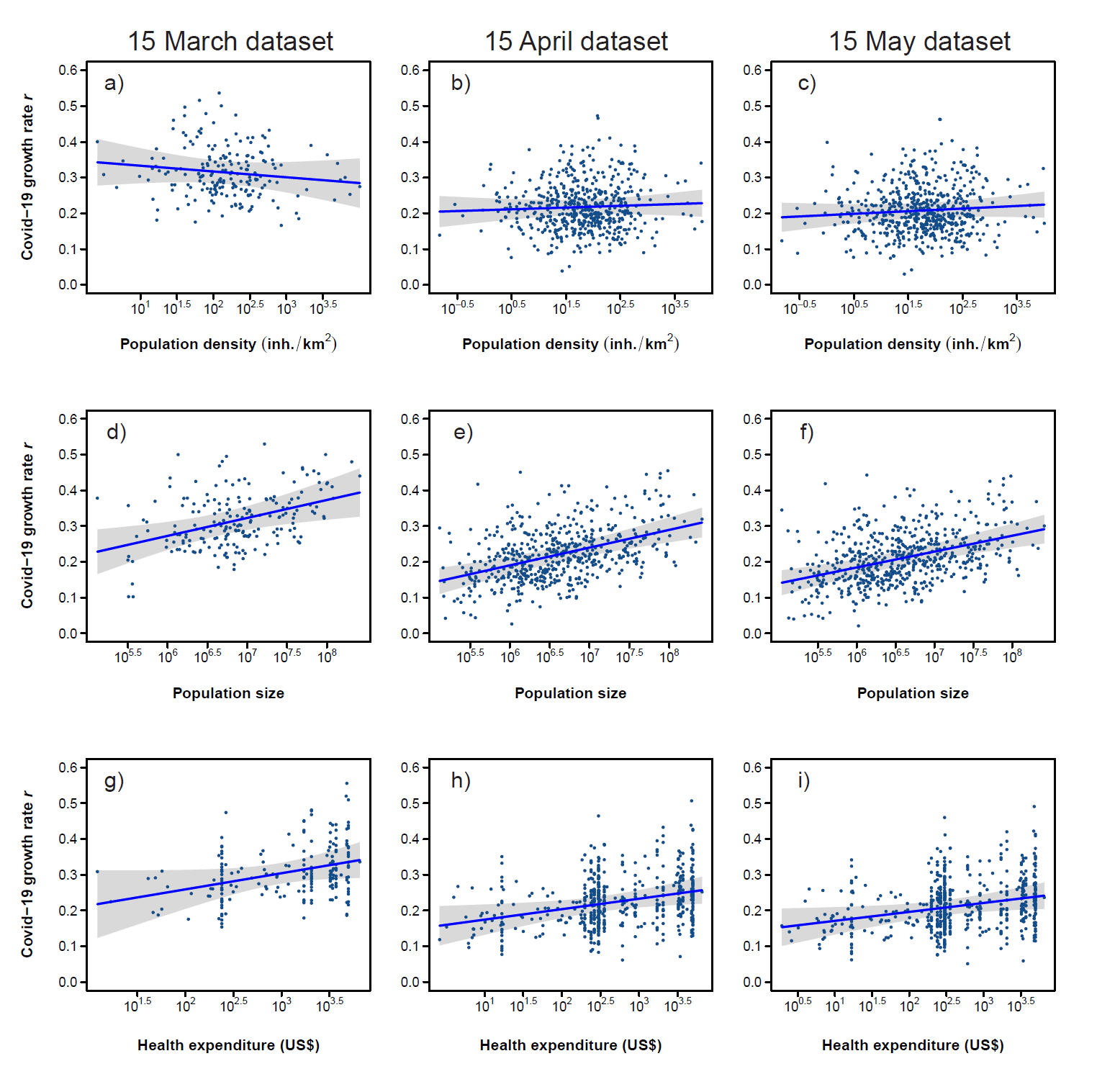

**Fig. S6.** Relationship between mean daily COVID-19 testing rate (mean daily testing rate up to May 31, 2020) and health expenditure (both variables log_10_-transformed) (Pearson’s correlation r = 0.79, *P* < 0.001, n = 84 countries). Data are reported in Dataset S2.

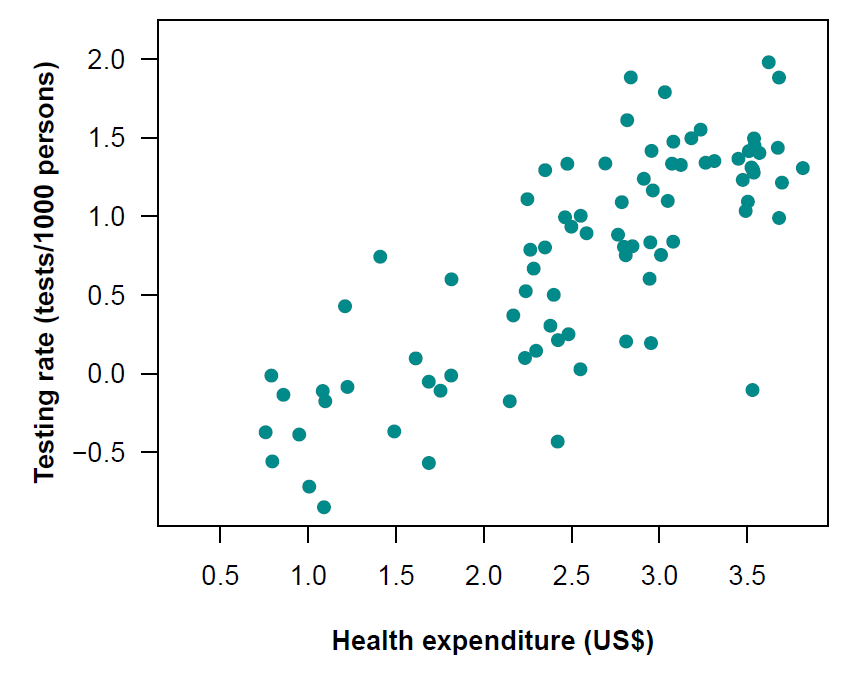

**Fig. S7.** Relationship between mean specific humidity and mean temperature (30-days period) of the outbreak regions (n = 586 regions). The Pearson’s correlation coefficient between temperature and log_10_-transformed humidity was r = 0.91. The pattern was identical if mean climate variables were calculated using a Δ14 days period (see Fig. S1).

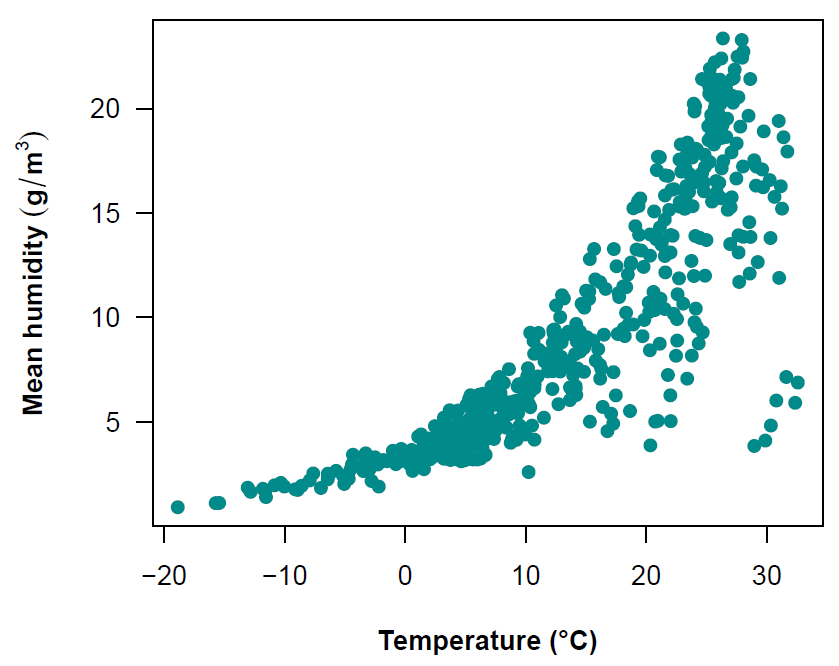

**Fig. S8.** Air transportation network among regions included in the dataset up to March 15, 2020 (n = 186; 9 regions not included in the network because of the lack of airports/connections), based on 40,050 direct flight connections among regions retrieved from the OpenFlights database.

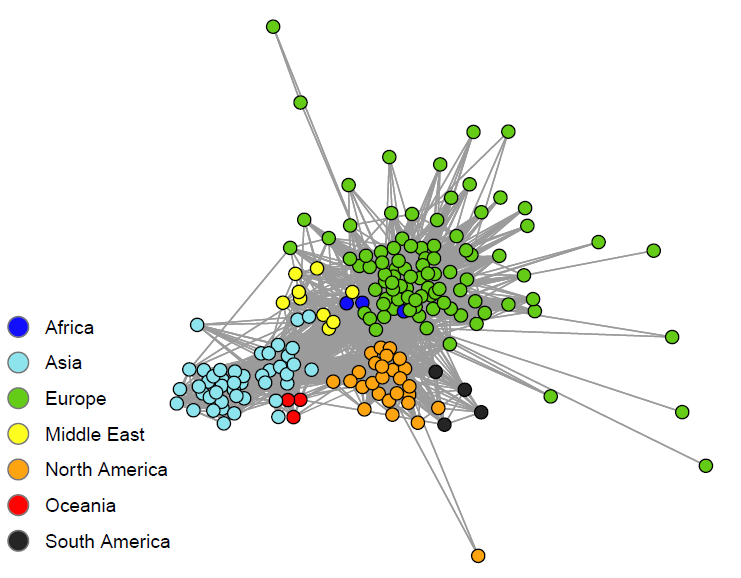

**Fig. S9.** Temporal variation of the relationships between independent variables and COVID-19 growth rate (standardized coefficients) (mixed models including humidity instead of temperature as a climatic predictor). We fitted regression models starting from regions experiencing outbreaks up to February 27, until we included all regions experiencing outbreaks up to May 31, 2020 (n = 586 regions). The plot includes humidity calculated using the 30-days period, but the pattern was identical when using the Δ14 days period (see Fig. S1). Shaded areas represent 95% confidence bands. When confidence bands do not cross the horizontal broken line (0 threshold), the effect of a given variable is statistically significant (*P* < 0.05).

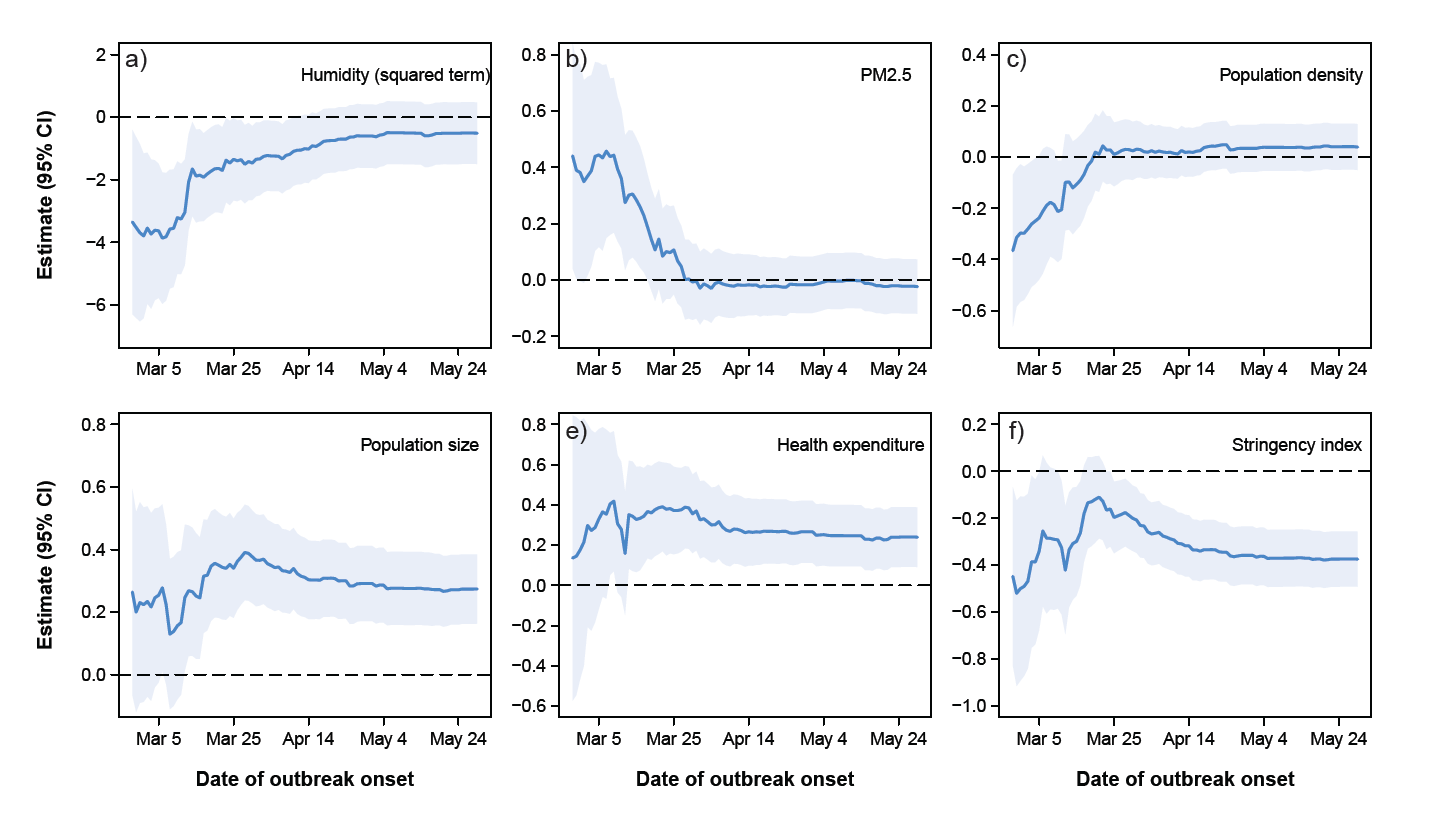

**Fig. S10.** Temporal variation of the relationships between independent variables and COVID-19 growth rate (standardized coefficients) (mixed models including temperature as a climatic predictor and population 65+ instead of health expenditure as socio-economic predictor). We fitted regression models starting from regions experiencing outbreaks up to February 27, until we included all regions experiencing outbreaks up to May 31, 2020 (n = 586 regions). The plot includes temperature calculated using the 30-days period, but the pattern was identical when using the Δ14 days period (see Fig. S1). Shaded areas represent 95% confidence bands. When confidence bands do not cross the horizontal broken line (0 threshold), the effect of a given variable is statistically significant (*P* < 0.05).

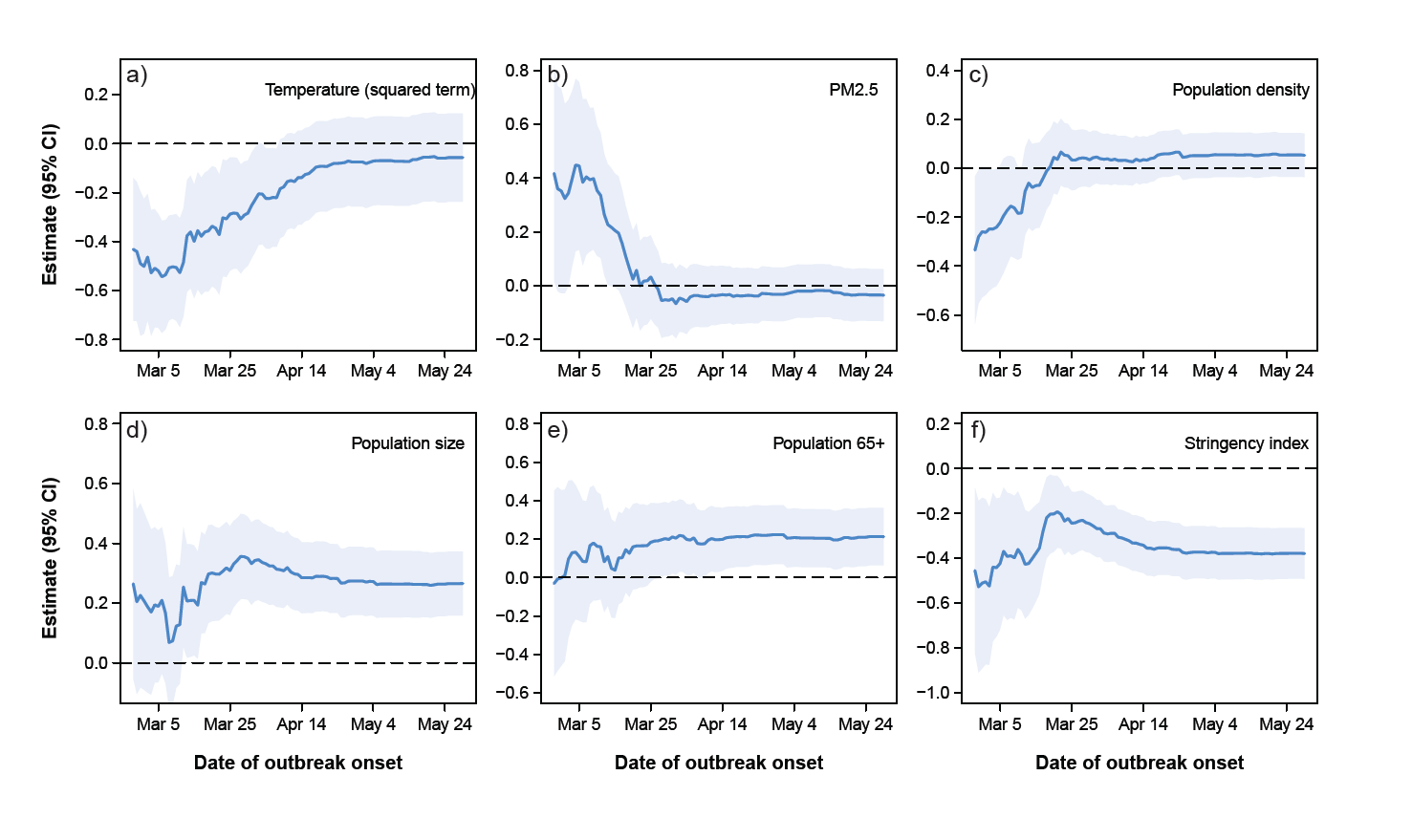

**Fig. S11.** Temporal variation of the relationships between independent variables and COVID-19 growth rate (standardized coefficients) (mixed models including humidity as a climatic predictor and population 65+ instead of health expenditure as socio-economic predictor). We fitted regression models starting from regions experiencing outbreaks up to February 27, until we included all regions experiencing outbreaks up to May 31, 2020 (n = 586 regions). The plot includes humidity calculated using the 30-days period, but the pattern was identical when using the Δ14 days period (see Fig. S1). Shaded areas represent 95% confidence bands. When confidence bands do not cross the horizontal broken line (0 threshold), the effect of a given variable is statistically significant (*P* < 0.05).

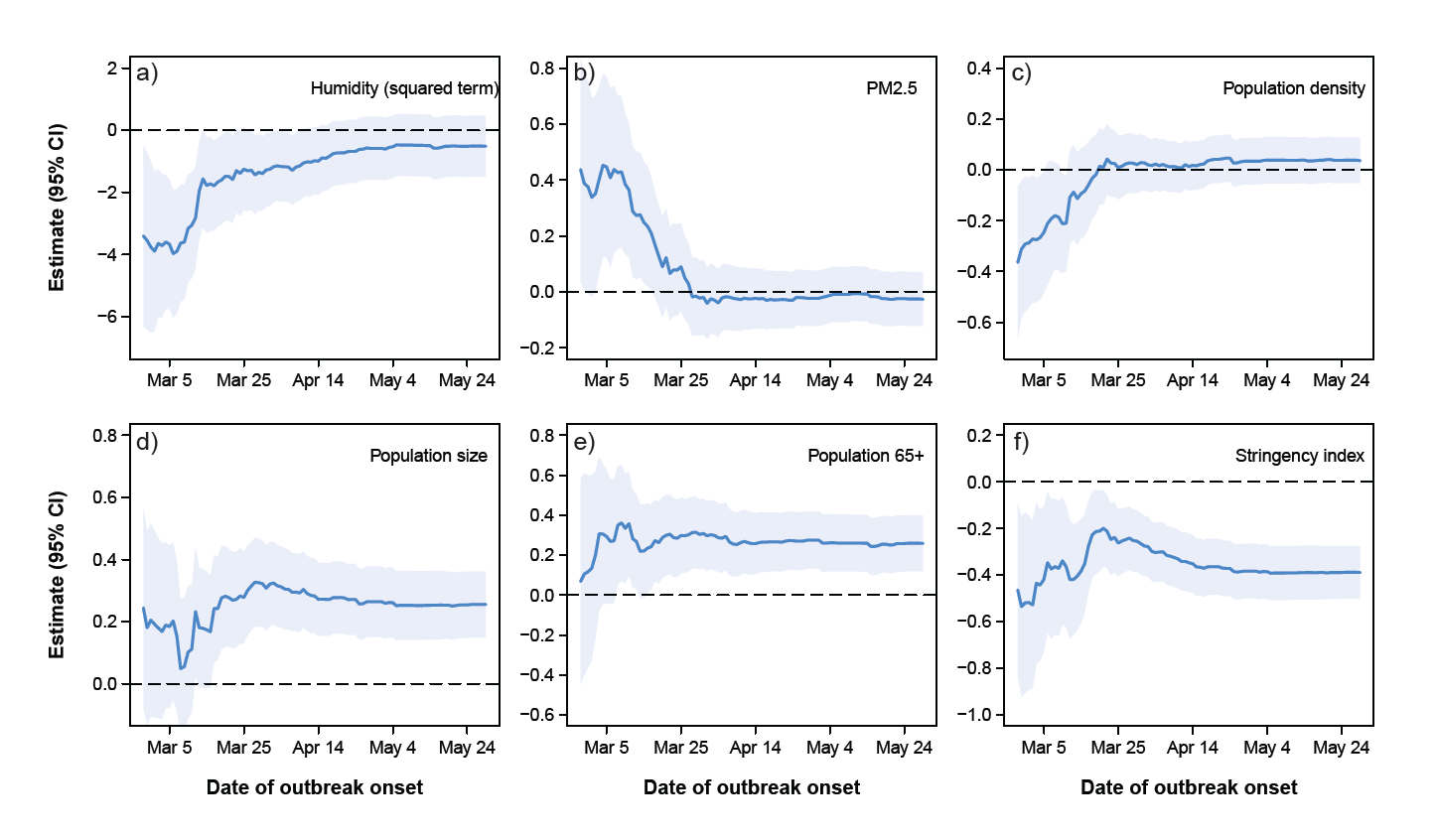

**Fig. S12.** Temporal variation of the relationships between independent variables and COVID-19 growth rate (standardized coefficients) (mixed models including maximum growth rate instead of mean daily growth rate as the dependent variable). We fitted regression models starting from regions experiencing outbreaks up to February 27, until we included all regions experiencing outbreaks up to May 31, 2020 (n = 586 regions). The plot includes temperature calculated using the 30-days period, but the pattern was identical when using the Δ14 days period (see Fig. S1). Shaded areas represent 95% confidence bands. When confidence bands do not cross the horizontal broken line (0 threshold), the effect of a given variable is statistically significant (P < 0.05).

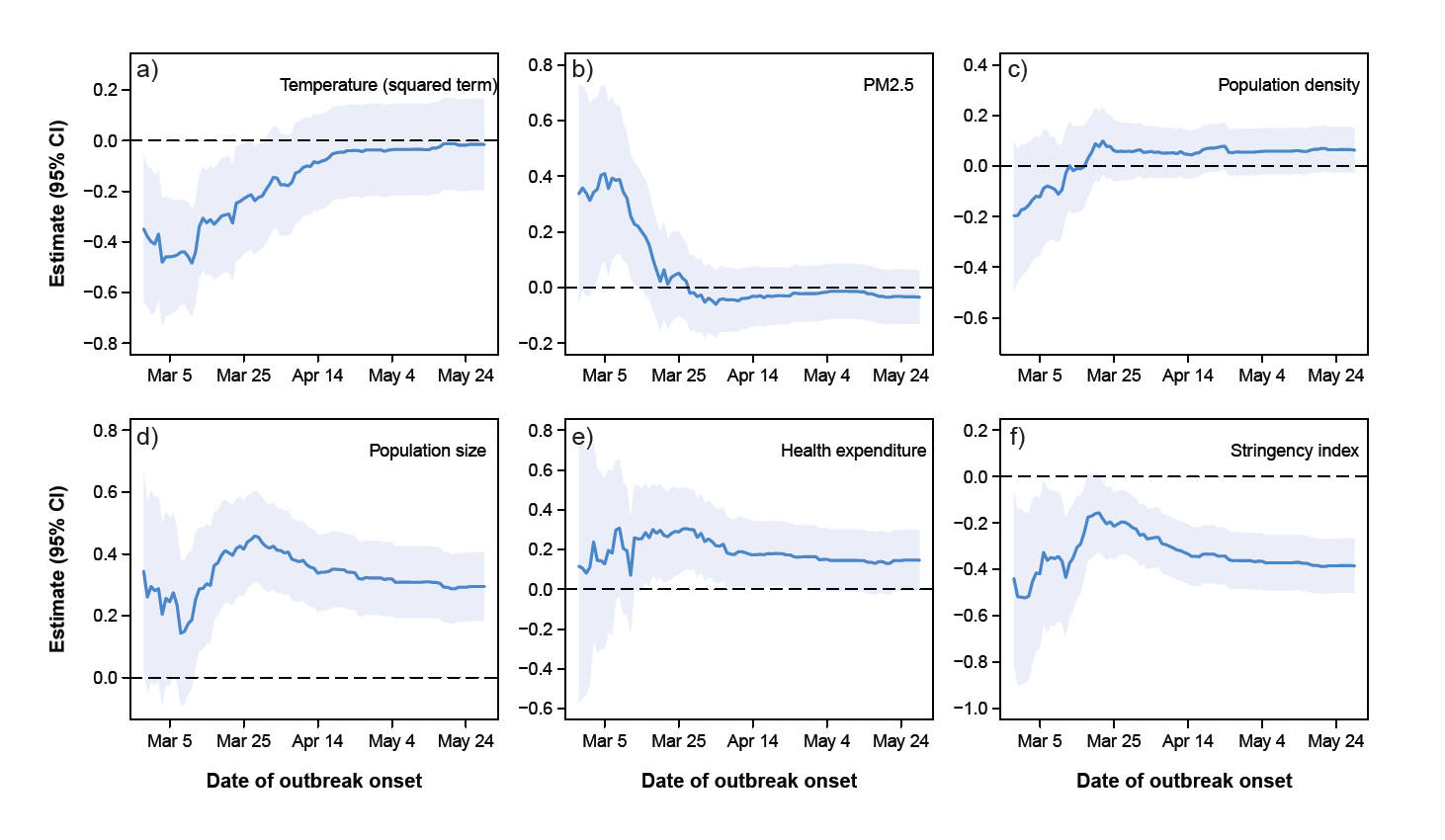

**Table S1.** Pearson’s correlation coefficients between candidate predictors of COVID-19 growth rates (n = 267 regions with local outbreaks occurring before March 21, 2020). Humidity, PM2.5, population density, population size, health expenditure, and region centrality were log_10_–transformed. Correlations refer to 30-days period temperature and humidity; the patterns were identical if a Δ14 days period was used (see Fig. S1). Sample size for correlations involving region centrality is 186 regions (only data up to March 15) (see Materials and Methods).

|  | Humidity | PM2.5 | Population  density | Population  size | Health  expenditure | Population  65+ | Region  centrality | Stringency index |
| --- | --- | --- | --- | --- | --- | --- | --- | --- |
| Temperature | 0.91 | -0.11 | 0.17 | 0.19 | -0.39 | -0.62 | 0.20 | 0.19 |
| Humidity |  | -0.13 | 0.23 | 0.13 | -0.31 | -0.31 | 0.20 | 0.15 |
| PM2.5 |  |  | 0.43 | 0.45 | -0.27 | -0.13 | 0.19 | 0.06 |
| Population density |  |  |  | 0.51 | -0.04 | -0.01 | 0.24 | -0.29 |
| Population size |  |  |  |  | -0.18 | -0.27 | 0.71 | -0.23 |
| Health expenditure |  |  |  |  |  | 0.63 | -0.20 | -0.49 |
| Population 65+ |  |  |  |  |  |  | -0.39 | -0.36 |
| Region centrality |  |  |  |  |  |  |  | 0.01 |

**Table S2.** Details on the variables included in the five alternative models shown in Fig. S4.

|  | Dependent variable | Independent variables |
| --- | --- | --- |
| a) | *r*_mean_ | Temperature + temperature^2^ + PM2.5 + population density + population size + health expenditure + stringency index |
| b) | *r*_mean_ | Temperature + temperature^2^ + PM2.5 + population density + population size + population 65+ + stringency index |
| c) | *r*_mean_ | Humidity + humidity^2^ + PM2.5 + population density + population size + health expenditure + stringency index |
| d) | *r*_mean_ | Humidity + humidity^2^ + PM2.5 + population density + population size + population 65+ + stringency index |
| e) | *r*_max_ | Temperature + temperature^2^ + PM2.5 + population density + population size + health expenditure + stringency index |

**Table S3a.** Parameter estimates for mixed models (LMMs) of COVID-19 growth rates including data up to March 15, April 15 or May 15, respectively, and temperature as a climatic predictor. Models include temperature calculated over the 30-days period (see Fig. S1). Estimates were obtained by Restricted Maximum Likelihood and denominator d.f. were estimated according to the Kenward-Roger method (Kuznetsova et al., 2017). LMMs included country identity as a random intercept effect to account for non-independence among regions belonging to the same country (e.g. US states, China provinces).

| Predictors | Estimate (s.e.) | *F* | d.f. | *P* |
| --- | --- | --- | --- | --- |
| a) *Data up to March 15 (n = 195 regions, 73 countries; AIC = -353.8)* | | | | |
| Temperature | 1.48 × 10^-3^ (1.37 × 10^-3^) | 1.15 | 1, 157 | 0.28 |
| Temperature^2^ | -2.39 × 10^-4^ (0.68 × 10^-4^) | 12.32 | 1, 184 | < 0.001 |
| PM2.5 | 8.51 × 10^-2^ (3.88 × 10^-2^) | 4.73 | 1, 181 | 0.031 |
| Population density | -1.62 × 10^-2^ (1.68 × 10^-2^) | 0.91 | 1, 187 | 0.34 |
| Population size | 5.02 × 10^-2^ (1.76 × 10^-2^) | 7.91 | 1, 166 | 0.006 |
| Health expenditure | 4.51 × 10^-2^ (2.44 × 10^-2^) | 3.39 | 1, 86 | 0.07 |
| Stringency index | -1.55 × 10^-3^ (0.59 × 10^-3^) | 6.54 | 1, 131 | 0.011 |
| Intercept | -1.63 × 10^-1^ (1.76 × 10^-1^) |  |  |  |
| b) *Data up to April 15 (n = 529 regions, 141 countries; AIC = -1111.9)* | | | | |
| Temperature | -0.12 × 10^-4^ (9.85 × 10^-4^) | 0.01 | 1, 483 | 0.99 |
| Temperature^2^ | -5.19 × 10^-5^ (3.71 × 10^-5^) | 1.95 | 1, 516 | 0.16 |
| PM2.5 | -0.85 × 10^-2^ (1.53 × 10^-2^) | 0.31 | 1, 516 | 0.58 |
| Population density | 4.87 × 10^-3^ (7.28 × 10^-3^) | 0.45 | 1, 508 | 0.50 |
| Population size | 4.96 × 10^-2^ (1.00 × 10^-2^) | 23.92 | 1, 512 | < 0.001 |
| Health expenditure | 2.92 × 10^-2^ (1.23 × 10^-2^) | 5.60 | 1, 221 | 0.019 |
| Stringency index | -1.36 × 10^-3^ (0.27 × 10^-3^) | 24.56 | 1, 460 | < 0.001 |
| Intercept | -8.45 × 10^-2^ (9.19 × 10^-2^) |  |  |  |
| c) *Data up to May 15 (n = 577 regions, 154 countries; AIC = -1222.4)* | | | | |
| Temperature | -4.99 × 10^-4^ (9.15 × 10^-4^) | 0.29 | 1, 522 | 0.59 |
| Temperature^2^ | -2.83 × 10^-5^ (3.54 × 10^-5^) | 0.63 | 1, 562 | 0.43 |
| PM2.5 | -0.66 × 10^-2^ (3.88 × 10^-2^) | 0.21 | 1, 568 | 0.65 |
| Population density | 7.31 × 10^-3^ (6.83 × 10^-3^) | 1.14 | 1, 551 | 0.28 |
| Population size | 4.44 × 10^-2^ (0.94 × 10^-2^) | 22.29 | 1, 559 | < 0.001 |
| Health expenditure | 2.50 × 10^-2^ (1.14 × 10^-2^) | 4.79 | 1, 241 | 0.029 |
| Stringency index | -1.52 × 10^-3^ (0.26 × 10^-3^) | 34.17 | 1, 514 | < 0.001 |
| Intercept | -3.64 × 10^-2^ (8.40 × 10^-2^) |  |  |  |

**Table S3b.** Parameter estimates for mixed models (LMMs) of COVID-19 growth rates including data up to March 15, April 15 or May 15, respectively, and temperature as a climatic predictor. Models include temperature calculated over the Δ14 days period (see Fig. S1). Estimates were obtained by Restricted Maximum Likelihood and denominator d.f. were estimated according to the Kenward-Roger method (Kuznetsova et al., 2017). LMMs included country identity as a random intercept effect to account for non-independence among regions belonging to the same country (e.g. US states, China provinces).

| Predictors | Estimate (s.e.) | *F* | d.f. | *P* |
| --- | --- | --- | --- | --- |
| a) *Data up to March 15 (n = 195 regions, 73 countries; AIC = -352.2)* | | | | |
| Temperature | 0.76 × 10^-3^ (1.32 × 10^-3^) | 0.32 | 1, 157 | 0.57 |
| Temperature^2^ | -2.14 × 10^-4^ (0.67 × 10^-4^) | 10.11 | 1, 186 | 0.002 |
| PM2.5 | 8.13 × 10^-2^ (3.92 × 10^-2^) | 4.20 | 1, 182 | 0.041 |
| Population density | -1.37 × 10^-2^ (1.68 × 10^-2^) | 0.65 | 1, 186 | 0.42 |
| Population size | 5.04 × 10^-2^ (1.77 × 10^-2^) | 7.87 | 1, 167 | 0.006 |
| Health expenditure | 4.33 × 10^-2^ (2.48 × 10^-2^) | 3.01 | 1, 87 | 0.09 |
| Stringency index | -1.61 × 10^-3^ (0.59 × 10^-3^) | 6.98 | 1, 133 | 0.009 |
| Intercept | -1.56 × 10^-1^ (1.77 × 10^-1^) |  |  |  |
| b) *Data up to April 15 (n = 529 regions, 141 countries; AIC = -1111.1)* | | | | |
| Temperature | -3.14 × 10^-4^ (8.89 × 10^-4^) | 0.12 | 1, 487 | 0.72 |
| Temperature^2^ | -3.67 × 10^-5^ (3.52 × 10^-5^) | 1.08 | 1, 515 | 0.30 |
| PM2.5 | -0.72 × 10^-2^ (1.53 × 10^-2^) | 0.22 | 1, 516 | 0.64 |
| Population density | 5.44 × 10^-3^ (7.27 × 10^-3^) | 0.56 | 1, 508 | 0.46 |
| Population size | 4.94 × 10^-2^ (1.01 × 10^-2^) | 23.57 | 1, 513 | < 0.001 |
| Health expenditure | 3.05 × 10^-2^ (1.23 × 10^-2^) | 6.11 | 1, 220 | 0.014 |
| Stringency index | -1.35 × 10^-3^ (0.27 × 10^-3^) | 24.32 | 1, 462 | < 0.001 |
| Intercept | -8.84 × 10^-2^ (9.19 × 10^-2^) |  |  |  |
| c) *Data up to May 15 (n = 577 regions, 154 countries; AIC = -1221.5)* | | | | |
| Temperature | -6.76 × 10^-4^ (8.58 × 10^-4^) | 0.62 | 1, 528 | 0.43 |
| Temperature^2^ | -1.51 × 10^-5^ (3.55 × 10^-5^) | 0.20 | 1, 561 | 0.65 |
| PM2.5 | -0.50 × 10^-2^ (1.43 × 10^-2^) | 0.12 | 1, 568 | 0.73 |
| Population density | 7.63 × 10^-3^ (6.85 × 10^-3^) | 1.24 | 1, 551 | 0.27 |
| Population size | 4.41 × 10^-2^ (0.93 × 10^-2^) | 21.93 | 1, 560 | < 0.001 |
| Health expenditure | 2.66 × 10^-2^ (1.14 × 10^-2^) | 5.44 | 1, 240 | 0.020 |
| Stringency index | -1.52 × 10^-3^ (0.26 × 10^-3^) | 34.05 | 1, 515 | < 0.001 |
| Intercept | -4.02 × 10^-2^ (8.40 × 10^-2^) |  |  |  |

**Table S4.** Parameter estimates for mixed models (LMMs) of COVID-19 growth rates including data up to March 15, April 15 or May 15, respectively, and humidity as a climatic predictor. Models include humidity calculated over the 30-days period ; the pattern was identical if the Δ14 days period was used (see Fig. S1). Estimates were obtained by Restricted Maximum Likelihood and denominator d.f. were estimated according to the Kenward-Roger method (Kuznetsova et al., 2017). LMMs included country identity as a random intercept effect to account for non-independence among regions belonging to the same country (e.g. US states, China provinces).

| Predictors | Estimate (s.e.) | *F* | d.f. | *P* |
| --- | --- | --- | --- | --- |
| a) *Data up to March 15 (n = 195 regions, 73 countries; AIC = -345.4)* | | | | |
| Humidity | -8.84 × 10^-1^ (3.64 × 10^-1^) | 5.83 | 1, 178 | 0.017 |
| Humidity^2^ | -1.95 × 10^-1^ (0.78 × 10^-1^) | 6.09 | 1, 184 | 0.014 |
| PM2.5 | 1.01 × 10^-1^ (0.39 × 10^-1^) | 6.31 | 1, 183 | 0.031 |
| Population density | -2.02 × 10^-2^ (1.72 × 10^-2^) | 1.36 | 1, 185 | 0.25 |
| Population size | 4.44 × 10^-2^ (1.80 × 10^-2^) | 5.88 | 1, 171 | 0.016 |
| Health expenditure | 6.11 × 10^-2^ (2.50 × 10^-2^) | 5.93 | 1, 86 | 0.017 |
| Stringency index | -1.56 × 10^-3^ (0.62 × 10^-3^) | 6.19 | 1, 142 | 0.014 |
| Intercept | -11.9 × 10^-1^ (4.35 × 10^-1^) |  |  |  |
| b) *Data up to April 15 (n = 529 regions, 141 countries; AIC = -1109.9)* | | | | |
| Humidity | -3.46 × 10^-1^ (2.05 × 10^-1^) | 2.84 | 1, 516 | 0.09 |
| Humidity^2^ | -7.78 × 10^-2^ (4.63 × 10^-2^) | 2.81 | 1, 508 | 0.09 |
| PM2.5 | -0.53 × 10^-2^ (1.53 × 10^-2^) | 0.12 | 1, 514 | 0.73 |
| Population density | 2.65 × 10^-3^ (7.25 × 10^-3^) | 0.13 | 1, 498 | 0.72 |
| Population size | 4.79 × 10^-2^ (1.01 × 10^-2^) | 22.31 | 1, 515 | < 0.001 |
| Health expenditure | 3.66 × 10^-2^ (1.18 × 10^-2^) | 9.60 | 1, 197 | 0.002 |
| Stringency index | -1.39 × 10^-3^ (0.27 × 10^-3^) | 25.35 | 1, 472 | < 0.001 |
| Intercept | -4.83 × 10^-1^ (2.36 × 10^-1^) |  |  |  |
| c) *Data up to May 15 (n = 577 regions, 154 countries; AIC = -1219.9)* | | | | |
| Humidity | -2.30 × 10^-1^ (1.91 × 10^-1^) | 1.43 | 1, 562 | 0.23 |
| Humidity^2^ | -5.08 × 10^-2^ (4.35 × 10^-2^) | 1.36 | 1, 552 | 0.24 |
| PM2.5 | -0.35 × 10^-2^ (1.43 × 10^-2^) | 0.06 | 1, 565 | 0.81 |
| Population density | 5.09 × 10^-3^ (6.84 × 10^-3^) | 0.55 | 1, 541 | 0.46 |
| Population size | 4.31 × 10^-2^ (0.94 × 10^-2^) | 21.04 | 1, 562 | < 0.001 |
| Health expenditure | 3.17 × 10^-2^ (1.08 × 10^-2^) | 8.56 | 1, 212 | 0.004 |
| Stringency index | -3.12 × 10^-1^ (2.18 × 10^-1^) | 35.34 | 1, 522 | < 0.001 |
| Intercept | -3.64 × 10^-2^ (8.40 × 10^-2^) |  |  |  |

**Table S5**. Parameter estimates for the mixed model (LMM) of COVID-19 growth rate including airport connections (region centrality) instead of population size, fitted on data up to March 15 (n = 186 regions from 69 countries). The model include temperature calculated over the 30-days period; the pattern was identical if the Δ14 days period was used (Fig. S1). Estimates were obtained by Restricted Maximum Likelihood and denominator d.f. were estimated according to the Kenward-Roger method (Kuznetsova et al., 2017). The model included country identity as a random intercept effect to account for non-independence among regions belonging to the same country (e.g. US states, China provinces).

| Predictors | Estimate (s.e.) | *F* | d.f. | *P* |
| --- | --- | --- | --- | --- |
| Temperature | 1.54 × 10^-3^ (1.39 × 10^-3^) | 1.21 | 1, 140 | 0.27 |
| Temperature^2^ | -2.45 × 10^-4^ (0.73 × 10^-4^) | 11.04 | 1, 177 | 0.001 |
| PM2.5 | 9.90 × 10^-2^ (4.09 × 10^-2^) | 5.75 | 1, 176 | 0.018 |
| Population density | -1.26 × 10^-2^ (1.96 × 10^-2^) | 0.41 | 1, 177 | 0.52 |
| Health expenditure | 1.74 × 10^-2^ (2.47 × 10^-2^) | 0.49 | 1, 77 | 0.49 |
| Region centrality | 0.73 × 10^-2^ (1.08 × 10^-2^) | 0.45 | 1, 176 | 0.50 |
| Stringency index | -2.12 × 10^-3^ (0.62 × 10^-3^) | 11.19 | 1, 153 | 0.001 |
| Intercept | 2.87 × 10^-1^ (1.04 × 10^-1^) |  |  |  |

**Table S6.** Sources of information used to obtain sub-national information on COVID-19 confirmed cases. Information on COVID-19 cases summarized in the links provided below were based on official reports from health institutions managing the COVID-19 outbreak. All websites were last accessed on June 25, 2020.

| Country | Source |
| --- | --- |
| Argentina (23 Provinces) | <https://datos.gob.ar/dataset/salud-covid-19-determinaciones-registradas-republica-argentina> |
| Brazil (27 States) | <https://github.com/wcota/covid19br> |
| Canada (Quebec only) | <https://en.wikipedia.org/wiki/2020_coronavirus_pandemic_in_Quebec> |
| Chile (16 Regions) | <https://github.com/MinCiencia/Datos-COVID19/tree/master/output/producto3> |
| Colombia (24 Departments) | <https://github.com/CovidDataProject/DataCovid19Colombia/tree/master/covid_19_data_colombia/covid_19_time_series> |
| Germany (16 Federal states) | <https://github.com/jgehrcke/covid-19-germany-gae/blob/master/cases-rki-by-state.csv> |
| India (29 States and Union territories) | <https://www.kaggle.com/sudalairajkumar/covid19-in-india> |
| Italy (20 Regions) | <https://raw.githubusercontent.com/pcm-dpc/COVID-19/master/dati-regioni/> |
| Japan (8 Regions) | <https://github.com/reustle/covid19japan-data/> |
| Mexico (32 Federal states) | <https://coronavirus.gob.mx/datos/#DownZCSV> |
| Peru (25 Regions) | <https://www.datosabiertos.gob.pe/dataset/casos-positivos-por-covid-19-ministerio-de-salud-minsa> |
| Russian Federation (83 Subjects) | <https://github.com/grwlf/COVID-19_plus_Russia> |
| South Africa (9 Provinces) | <https://raw.githubusercontent.com/dsfsi/covid19za/master/data/covid19za_provincial_cumulative_timeline_confirmed.csv> |
| Spain (18 Autonomous communities) | <https://cnecovid.isciii.es/covid19/> |
| Sweden (21 Counties) | <https://raw.githubusercontent.com/jannesgg/swe-covid-19/master/time_series_confimed-confirmed.csv> |
| United Kingdom (9 Regions, 3 Countries) | <https://coronavirus.data.gov.uk/> (England) and <https://github.com/tomwhite/covid-19-uk-data/tree/master/data> (N. Ireland, Scotland, Wales) |
| United States (55 States and Territories) | [https://github.com/CSSEGISandData/COVID-19/blob/master/csse_covid_19_data/](https://github.com/CSSEGISandData/COVID-19/blob/master/csse_covid_19_data/csse_covid_19_time_series/time_series_covid19_confirmed_US.csv) |

**Table S7.** List of country/region data included in the analyses and estimates of daily confirmed COVID-19 cases growth rates (*r* = mean daily growth rate during the exponential phase, *r*_max_ = maximum daily growth rates), calculated when at least 10 days of data were available after a selected minimum threshold of 25 cases was reached (n = 586 regions experiencing local outbreaks up to May 31, 2020, i.e. day 152). The day when 25 cases were reached (day of outbreak onset) is also reported (day 1 = January 1).

| Country | Region | Day of outbreak onset | *r*_mean_ | *r*_max_ |
| --- | --- | --- | --- | --- |
| Afghanistan | Afghanistan | 82 | 0.214 | 0.245 |
| Albania | Albania | 73 | 0.123 | 0.166 |
| Algeria | Algeria | 73 | 0.197 | 0.250 |
| Angola | Angola | 113 | 0.035 | 0.085 |
| Argentina | Argentina_Buenos Aires | 71 | 0.231 | 0.238 |
| Argentina | Argentina_Chaco | 90 | 0.127 | 0.111 |
| Argentina | Argentina_Cordoba | 85 | 0.180 | 0.197 |
| Argentina | Argentina_Corrientes | 109 | 0.059 | 0.114 |
| Argentina | Argentina_Entre Rios | 100 | 0.028 | 0.082 |
| Argentina | Argentina_La Rioja | 104 | 0.061 | 0.094 |
| Argentina | Argentina_Mendoza | 94 | 0.110 | 0.142 |
| Argentina | Argentina_Misiones | 150 | 0.040 | 0.052 |
| Argentina | Argentina_Neuquen | 90 | 0.126 | 0.164 |
| Argentina | Argentina_Rio Negro | 99 | 0.220 | 0.233 |
| Argentina | Argentina_Santa Cruz | 107 | 0.065 | 0.072 |
| Argentina | Argentina_Santa Fe | 84 | 0.200 | 0.196 |
| Argentina | Argentina_Santiago del Estero | 139 | 0.009 | 0.009 |
| Argentina | Argentina_Tierra del Fuego | 93 | 0.054 | 0.080 |
| Argentina | Argentina_Tucuman | 116 | 0.050 | 0.057 |
| Armenia | Armenia | 75 | 0.372 | 0.345 |
| Australia | Australia_Australian Capital Territory | 83 | 0.146 | 0.153 |
| Australia | Australia_New South Wales | 66 | 0.198 | 0.209 |
| Australia | Australia_Northern Territory | 95 | 0.019 | 0.018 |
| Australia | Australia_Queensland | 73 | 0.224 | 0.275 |
| Australia | Australia_South Australia | 76 | 0.254 | 0.296 |
| Australia | Australia_Tasmania | 83 | 0.199 | 0.186 |
| Australia | Australia_Victoria | 73 | 0.230 | 0.323 |
| Australia | Australia_Western Australia | 76 | 0.271 | 0.301 |
| Austria | Austria | 64 | 0.308 | 0.305 |
| Azerbaijan | Azerbaijan | 76 | 0.183 | 0.187 |
| Bahamas | Bahamas | 95 | 0.095 | 0.108 |
| Bahrain | Bahrain | 57 | 0.165 | 0.238 |
| Bangladesh | Bangladesh | 81 | 0.229 | 0.317 |
| Belarus | Belarus | 73 | 0.171 | 0.276 |
| Belgium | Belgium | 65 | 0.391 | 0.374 |
| Benin | Benin | 97 | 0.062 | 0.112 |
| Bhutan | Bhutan | 146 | 0.083 | 0.110 |
| Bolivia | Bolivia | 83 | 0.205 | 0.229 |
| Bosnia and Herzegovina | Bosnia and Herzegovina | 76 | 0.263 | 0.308 |
| Botswana | Botswana | 138 | 0.032 | 0.023 |
| Brazil | Brazil_Acre | 87 | 0.089 | 0.083 |
| Brazil | Brazil_Alagoas | 96 | 0.180 | 0.220 |
| Brazil | Brazil_Amapa | 95 | 0.329 | 0.358 |
| Brazil | Brazil_Amazonas | 82 | 0.202 | 0.190 |
| Brazil | Brazil_Bahia | 78 | 0.187 | 0.212 |
| Brazil | Brazil_Ceara | 80 | 0.250 | 0.267 |
| Brazil | Brazil_Distrito Federal | 78 | 0.309 | 0.342 |
| Brazil | Brazil_Espirito Santo | 81 | 0.137 | 0.182 |
| Brazil | Brazil_Goias | 84 | 0.165 | 0.165 |
| Brazil | Brazil_Maranhao | 90 | 0.245 | 0.221 |
| Brazil | Brazil_Mato Grosso | 91 | 0.219 | 0.220 |
| Brazil | Brazil_Mato Grosso do Sul | 86 | 0.130 | 0.136 |
| Brazil | Brazil_Minas Gerais | 79 | 0.371 | 0.375 |
| Brazil | Brazil_Para | 90 | 0.230 | 0.237 |
| Brazil | Brazil_Paraiba | 93 | 0.177 | 0.237 |
| Brazil | Brazil_Parana | 80 | 0.159 | 0.146 |
| Brazil | Brazil_Pernambuco | 79 | 0.178 | 0.257 |
| Brazil | Brazil_Piaui | 97 | 0.131 | 0.176 |
| Brazil | Brazil_Rio de Janeiro | 76 | 0.288 | 0.300 |
| Brazil | Brazil_Rio Grande do Norte | 87 | 0.241 | 0.244 |
| Brazil | Brazil_Rio Grande do Sul | 79 | 0.283 | 0.284 |
| Brazil | Brazil_Rondonia | 99 | 0.176 | 0.206 |
| Brazil | Brazil_Roraima | 92 | 0.128 | 0.168 |
| Brazil | Brazil_Santa Catarina | 80 | 0.251 | 0.242 |
| Brazil | Brazil_Sao Paulo | 71 | 0.325 | 0.329 |
| Brazil | Brazil_Sergipe | 94 | 0.153 | 0.226 |
| Brazil | Brazil_Tocantins | 102 | 0.165 | 0.221 |
| Brunei | Brunei | 73 | 0.122 | 0.118 |
| Bulgaria | Bulgaria | 74 | 0.194 | 0.219 |
| Burkina Faso | Burkina Faso | 79 | 0.275 | 0.283 |
| Burma | Burma | 101 | 0.212 | 0.218 |
| Burundi | Burundi | 139 | 0.101 | 0.104 |
| Cabo Verde | Cabo Verde | 106 | 0.069 | 0.104 |
| Cambodia | Cambodia | 77 | 0.182 | 0.201 |
| Cameroon | Cameroon | 81 | 0.213 | 0.306 |
| Canada | Canada_Alberta | 73 | 0.302 | 0.306 |
| Canada | Canada_British Columbia | 68 | 0.247 | 0.321 |
| Canada | Canada_Manitoba | 85 | 0.245 | 0.257 |
| Canada | Canada_New Brunswick | 86 | 0.181 | 0.183 |
| Canada | Canada_Newfoundland and Labrador | 84 | 0.308 | 0.353 |
| Canada | Canada_Nova Scotia | 82 | 0.240 | 0.242 |
| Canada | Canada_Ontario | 66 | 0.265 | 0.322 |
| Canada | Canada_Prince Edward Island | 99 | 0.004 | 0.003 |
| Canada | Canada_Quebec S | 75 | 0.414 | 0.598 |
| Canada | Canada_Saskatchewan | 81 | 0.255 | 0.236 |
| Central African Republic | Central African Republic | 119 | 0.092 | 0.210 |
| Chad | Chad | 107 | 0.133 | 0.173 |
| Chile | Chile_Antofagasta | 88 | 0.114 | 0.140 |
| Chile | Chile_Araucania | 82 | 0.353 | 0.344 |
| Chile | Chile_Arica y Parinacota | 96 | 0.246 | 0.240 |
| Chile | Chile_Atacama | 117 | 0.100 | 0.130 |
| Chile | Chile_Biobio | 81 | 0.258 | 0.283 |
| Chile | Chile_Coquimbo | 90 | 0.116 | 0.128 |
| Chile | Chile_Los Lagos | 83 | 0.212 | 0.217 |
| Chile | Chile_Los Rios | 88 | 0.216 | 0.210 |
| Chile | Chile_Magallanes | 88 | 0.299 | 0.337 |
| Chile | Chile_Maule | 82 | 0.077 | 0.188 |
| Chile | Chile_Metropolitana | 73 | 0.414 | 0.444 |
| Chile | Chile_Nuble | 77 | 0.227 | 0.241 |
| Chile | Chile_O'Higgins | 92 | 0.057 | 0.049 |
| Chile | Chile_Tarapaca | 99 | 0.145 | 0.145 |
| Chile | Chile_Valparaiso | 84 | 0.261 | 0.251 |
| China | China_Anhui | 25 | 0.327 | 0.325 |
| China | China_Beijing | 24 | 0.232 | 0.252 |
| China | China_Chongqing | 24 | 0.397 | 0.383 |
| China | China_Fujian | 26 | 0.265 | 0.247 |
| China | China_Gansu | 30 | 0.187 | 0.206 |
| China | China_Guangdong | 22 | 0.352 | 0.369 |
| China | China_Guangxi | 26 | 0.170 | 0.170 |
| China | China_Guizhou | 31 | 0.198 | 0.201 |
| China | China_Hainan | 27 | 0.108 | 0.133 |
| China | China_Hebei | 28 | 0.267 | 0.267 |
| China | China_Heilongjiang | 28 | 0.232 | 0.240 |
| China | China_Henan | 25 | 0.466 | 0.443 |
| China | China_Hong Kong | 38 | 0.163 | 0.183 |
| China | China_Hunan | 25 | 0.409 | 0.400 |
| China | China_Inner Mongolia | 33 | 0.133 | 0.128 |
| China | China_Jiangsu | 26 | 0.341 | 0.347 |
| China | China_Jiangxi | 26 | 0.376 | 0.342 |
| China | China_Jilin | 34 | 0.185 | 0.182 |
| China | China_Liaoning | 27 | 0.144 | 0.146 |
| China | China_Ningxia | 32 | 0.072 | 0.087 |
| China | China_Shaanxi | 27 | 0.228 | 0.214 |
| China | China_Shandong | 25 | 0.393 | 0.387 |
| China | China_Shanghai | 25 | 0.244 | 0.256 |
| China | China_Shanxi | 28 | 0.202 | 0.204 |
| China | China_Sichuan | 25 | 0.337 | 0.342 |
| China | China_Tianjin | 29 | 0.154 | 0.186 |
| China | China_Xinjiang | 35 | 0.093 | 0.094 |
| China | China_Yunnan | 27 | 0.290 | 0.279 |
| China | China_Zhejiang | 23 | 0.389 | 0.400 |
| Colombia | Colombia_Amazonas | 116 | 0.239 | 0.287 |
| Colombia | Colombia_Antioquia | 81 | 0.219 | 0.230 |
| Colombia | Colombia_Atlantico | 90 | 0.091 | 0.161 |
| Colombia | Colombia_Bolivar | 85 | 0.114 | 0.164 |
| Colombia | Colombia_Boyaca | 101 | 0.062 | 0.087 |
| Colombia | Colombia_Caldas | 101 | 0.052 | 0.036 |
| Colombia | Colombia_Casanare | 134 | 0.060 | 0.068 |
| Colombia | Colombia_Cauca | 113 | 0.065 | 0.073 |
| Colombia | Colombia_Cesar | 102 | 0.116 | 0.195 |
| Colombia | Colombia_Choco | 126 | 0.113 | 0.116 |
| Colombia | Colombia_Cordoba | 119 | 0.099 | 0.157 |
| Colombia | Colombia_Cundinamarca | 85 | 0.110 | 0.129 |
| Colombia | Colombia_Huila | 92 | 0.055 | 0.065 |
| Colombia | Colombia_La Guajira | 131 | 0.064 | 0.147 |
| Colombia | Colombia_Magdalena | 99 | 0.213 | 0.235 |
| Colombia | Colombia_Meta | 75 | 0.190 | 0.193 |
| Colombia | Colombia_Narino | 100 | 0.091 | 0.122 |
| Colombia | Colombia_Norte de Santander | 94 | 0.099 | 0.121 |
| Colombia | Colombia_Quindio | 99 | 0.024 | 0.050 |
| Colombia | Colombia_Risaralda | 89 | 0.103 | 0.171 |
| Colombia | Colombia_Santander | 102 | 0.079 | 0.145 |
| Colombia | Colombia_Tolima | 104 | 0.076 | 0.127 |
| Colombia | Colombia_Valle del Cauca | 81 | 0.245 | 0.270 |
| Comoros | Comoros | 141 | 0.235 | 0.271 |
| Congo (Brazzaville) | Congo (Brazzaville) | 96 | 0.174 | 0.240 |
| Congo (Kinshasa) | Congo (Kinshasa) | 82 | 0.121 | 0.174 |
| Costa Rica | Costa Rica | 74 | 0.244 | 0.267 |
| Cote d'Ivoire | Cote d'Ivoire | 83 | 0.349 | 0.307 |
| Croatia | Croatia | 73 | 0.233 | 0.283 |
| Cuba | Cuba | 82 | 0.207 | 0.230 |
| Cyprus | Cyprus | 74 | 0.237 | 0.229 |
| Czechia | Czechia | 68 | 0.379 | 0.386 |
| Denmark | Denmark | 68 | 0.717 | 0.732 |
| Djibouti | Djibouti | 91 | 0.192 | 0.194 |
| Dominican Republic | Dominican Republic | 79 | 0.494 | 0.498 |
| Ecuador | Ecuador | 74 | 0.574 | 0.582 |
| Egypt | Egypt | 68 | 0.197 | 0.211 |
| El Salvador | El Salvador | 90 | 0.132 | 0.163 |
| Equatorial Guinea | Equatorial Guinea | 105 | 0.169 | 0.285 |
| Estonia | Estonia | 73 | 0.262 | 0.267 |
| Eswatini | Eswatini | 112 | 0.148 | 0.174 |
| Ethiopia | Ethiopia | 91 | 0.061 | 0.121 |
| Finland | Finland | 69 | 0.432 | 0.442 |
| France | France | 58 | 0.358 | 0.334 |
| France | France_French Guiana | 85 | 0.150 | 0.163 |
| France | France_French Polynesia | 84 | 0.045 | 0.042 |
| France | France_Guadeloupe | 78 | 0.191 | 0.200 |
| France | France_Martinique | 80 | 0.145 | 0.158 |
| France | France_Reunion | 80 | 0.303 | 0.288 |
| Gabon | Gabon | 98 | 0.100 | 0.146 |
| Gambia | Gambia | 143 | 0.011 | 0.019 |
| Georgia | Georgia | 73 | 0.084 | 0.119 |
| Germany | Germany_Baden-Wurttemberg | 62 | 0.432 | 0.429 |
| Germany | Germany_Bavaria | 62 | 0.297 | 0.315 |
| Germany | Germany_Berlin | 67 | 0.420 | 0.430 |
| Germany | Germany_Brandenburg | 71 | 0.333 | 0.317 |
| Germany | Germany_Hamburg | 68 | 0.425 | 0.422 |
| Germany | Germany_Hesse | 67 | 0.405 | 0.398 |
| Germany | Germany_Lower Saxony | 66 | 0.362 | 0.432 |
| Germany | Germany_Mecklenburg-Western Pomerania | 72 | 0.285 | 0.366 |
| Germany | Germany_North Rhine-Westphalia | 59 | 0.446 | 0.468 |
| Germany | Germany_Rhineland-Palatinate | 70 | 0.461 | 0.472 |
| Germany | Germany_Saarland | 72 | 0.298 | 0.285 |
| Germany | Germany_Saxony | 70 | 0.357 | 0.366 |
| Germany | Germany_Saxony-Anhalt | 72 | 0.278 | 0.337 |
| Germany | Germany_Schleswig-Holstein | 71 | 0.304 | 0.308 |
| Germany | Germany_Thuringen | 73 | 0.277 | 0.330 |
| Ghana | Ghana | 83 | 0.406 | 0.416 |
| Greece | Greece | 65 | 0.216 | 0.325 |
| Guatemala | Guatemala | 86 | 0.112 | 0.147 |
| Guinea | Guinea | 92 | 0.349 | 0.355 |
| Guinea-Bissau | Guinea-Bissau | 98 | 0.274 | 0.374 |
| Guyana | Guyana | 97 | 0.067 | 0.083 |
| Haiti | Haiti | 98 | 0.105 | 0.172 |
| Honduras | Honduras | 82 | 0.260 | 0.276 |
| Hungary | Hungary | 74 | 0.196 | 0.199 |
| Iceland | Iceland | 64 | 0.171 | 0.170 |
| India | India_Andhra Pradesh | 91 | 0.348 | 0.325 |
| India | India_Assam | 96 | 0.166 | 0.223 |
| India | India_Bihar | 94 | 0.105 | 0.169 |
| India | India_Chhattisgarh | 103 | 0.123 | 0.197 |
| India | India_Delhi | 81 | 0.314 | 0.330 |
| India | India_Goa | 139 | 0.191 | 0.334 |
| India | India_Gujarat | 83 | 0.166 | 0.279 |
| India | India_Haryana | 83 | 0.196 | 0.257 |
| India | India_Himachal Pradesh | 101 | 0.095 | 0.164 |
| India | India_Jammu and Kashmir | 86 | 0.237 | 0.251 |
| India | India_Jharkhand | 106 | 0.110 | 0.146 |
| India | India_Karnataka | 82 | 0.187 | 0.172 |
| India | India_Kerala | 77 | 0.305 | 0.296 |
| India | India_Madhya Pradesh | 88 | 0.298 | 0.273 |
| India | India_Maharashtra | 75 | 0.163 | 0.233 |
| India | India_Manipur | 142 | 0.149 | 0.222 |
| India | India_Odisha | 98 | 0.089 | 0.172 |
| India | India_Punjab | 84 | 0.124 | 0.331 |
| India | India_Rajasthan | 83 | 0.198 | 0.248 |
| India | India_Tamil Nadu | 86 | 0.341 | 0.403 |
| India | India_Telengana | 83 | 0.198 | 0.273 |
| India | India_Tripura | 126 | 0.274 | 0.293 |
| India | India_Uttar Pradesh | 82 | 0.171 | 0.184 |
| India | India_Uttarakhand | 97 | 0.136 | 0.230 |
| India | India_West Bengal | 91 | 0.244 | 0.248 |
| Indonesia | Indonesia | 70 | 0.309 | 0.351 |
| Iran | Iran | 53 | 0.442 | 0.463 |
| Iraq | Iraq | 62 | 0.119 | 0.132 |
| Ireland | Ireland | 70 | 0.346 | 0.371 |
| Israel | Israel | 66 | 0.235 | 0.302 |
| Italy | Italy_Abruzzo | 69 | 0.263 | 0.284 |
| Italy | Italy_Basilicata | 78 | 0.275 | 0.278 |
| Italy | Italy_Calabria | 72 | 0.248 | 0.254 |
| Italy | Italy_Campania | 63 | 0.271 | 0.265 |
| Italy | Italy_Emilia-Romagna | 56 | 0.530 | 0.537 |
| Italy | Italy_Friuli Venezia Giulia | 66 | 0.330 | 0.343 |
| Italy | Italy_Lazio | 64 | 0.233 | 0.286 |
| Italy | Italy_Liguria | 63 | 0.323 | 0.348 |
| Italy | Italy_Lombardia | 53 | 0.386 | 0.385 |
| Italy | Italy_Marche | 61 | 0.400 | 0.408 |
| Italy | Italy_Molise | 77 | 0.223 | 0.236 |
| Italy | Italy_Piemonte | 61 | 0.367 | 0.355 |
| Italy | Italy_Puglia | 67 | 0.272 | 0.258 |
| Italy | Italy_Sardegna | 71 | 0.255 | 0.264 |
| Italy | Italy_Sicilia | 67 | 0.214 | 0.224 |
| Italy | Italy_Toscana | 64 | 0.369 | 0.357 |
| Italy | Italy_Trentino | 68 | 0.472 | 0.506 |
| Italy | Italy_Umbria | 68 | 0.272 | 0.278 |
| Italy | Italy_Valle d'Aosta | 72 | 0.395 | 0.408 |
| Italy | Italy_Veneto | 54 | 0.360 | 0.399 |
| Jamaica | Jamaica | 85 | 0.082 | 0.177 |
| Japan | Japan_Chubu | 57 | 0.141 | 0.202 |
| Japan | Japan_Chugoku | 94 | 0.166 | 0.218 |
| Japan | Japan_Hokkaido | 55 | 0.146 | 0.149 |
| Japan | Japan_Kansai | 65 | 0.214 | 0.189 |
| Japan | Japan_Kanto | 51 | 0.106 | 0.249 |
| Japan | Japan_Kyushu | 80 | 0.189 | 0.239 |
| Japan | Japan_Shikoku | 92 | 0.138 | 0.140 |
| Japan | Japan_Tohoku | 92 | 0.194 | 0.200 |
| Jordan | Jordan | 77 | 0.229 | 0.232 |
| Kazakhstan | Kazakhstan | 77 | 0.222 | 0.323 |
| Kenya | Kenya | 84 | 0.196 | 0.229 |
| Korea, South | Korea, South | 40 | 0.593 | 0.669 |
| Kosovo | Kosovo | 86 | 0.077 | 0.118 |
| Kuwait | Kuwait | 57 | 0.082 | 0.162 |
| Kyrgyzstan | Kyrgyzstan | 84 | 0.190 | 0.189 |
| Latvia | Latvia | 74 | 0.262 | 0.277 |
| Lebanon | Lebanon | 68 | 0.220 | 0.215 |
| Liberia | Liberia | 99 | 0.161 | 0.159 |
| Libya | Libya | 103 | 0.168 | 0.196 |
| Lithuania | Lithuania | 77 | 0.378 | 0.404 |
| Luxembourg | Luxembourg | 73 | 0.375 | 0.437 |
| Madagascar | Madagascar | 87 | 0.196 | 0.207 |
| Malawi | Malawi | 114 | 0.175 | 0.303 |
| Malaysia | Malaysia | 60 | 0.198 | 0.323 |
| Mali | Mali | 90 | 0.110 | 0.151 |
| Mauritania | Mauritania | 136 | 0.377 | 0.372 |
| Mauritius | Mauritius | 82 | 0.242 | 0.258 |
| Mexico | Mexico_Aguascalientes | 85 | 0.056 | 0.042 |
| Mexico | Mexico_Baja California | 82 | 0.212 | 0.261 |
| Mexico | Mexico_Baja California Sur | 90 | 0.169 | 0.167 |
| Mexico | Mexico_Campeche | 102 | 0.093 | 0.094 |
| Mexico | Mexico_Chiapas | 94 | 0.080 | 0.079 |
| Mexico | Mexico_Chihuahua | 90 | 0.161 | 0.159 |
| Mexico | Mexico_Coahuila | 86 | 0.222 | 0.254 |
| Mexico | Mexico_Colima | 115 | 0.068 | 0.105 |
| Mexico | Mexico_Distrito Federal | 72 | 0.281 | 0.265 |
| Mexico | Mexico_Durango | 110 | 0.076 | 0.091 |
| Mexico | Mexico_Guanajuato | 83 | 0.069 | 0.059 |
| Mexico | Mexico_Guerrero | 88 | 0.094 | 0.137 |
| Mexico | Mexico_Hidalgo | 87 | 0.152 | 0.164 |
| Mexico | Mexico_Jalisco | 76 | 0.225 | 0.241 |
| Mexico | Mexico_Mexico | 73 | 0.154 | 0.143 |
| Mexico | Mexico_Michoacan | 89 | 0.088 | 0.111 |
| Mexico | Mexico_Morelos | 93 | 0.127 | 0.138 |
| Mexico | Mexico_Nayarit | 100 | 0.083 | 0.078 |
| Mexico | Mexico_Nuevo Leon | 77 | 0.169 | 0.170 |
| Mexico | Mexico_Oaxaca | 91 | 0.080 | 0.120 |
| Mexico | Mexico_Puebla | 80 | 0.172 | 0.200 |
| Mexico | Mexico_Queretaro | 86 | 0.057 | 0.100 |
| Mexico | Mexico_Quintana Roo | 81 | 0.182 | 0.199 |
| Mexico | Mexico_San Luis Potosi | 87 | 0.058 | 0.088 |
| Mexico | Mexico_Sinaloa | 86 | 0.176 | 0.202 |
| Mexico | Mexico_Sonora | 90 | 0.120 | 0.146 |
| Mexico | Mexico_Tabasco | 85 | 0.164 | 0.163 |
| Mexico | Mexico_Tamaulipas | 92 | 0.118 | 0.155 |
| Mexico | Mexico_Tlaxcala | 97 | 0.133 | 0.125 |
| Mexico | Mexico_Veracruz | 86 | 0.103 | 0.142 |
| Mexico | Mexico_Yucatan | 82 | 0.110 | 0.116 |
| Mexico | Mexico_Zacatecas | 106 | 0.103 | 0.110 |
| Moldova | Moldova | 77 | 0.228 | 0.256 |
| Mongolia | Mongolia | 105 | 0.234 | 0.318 |
| Montenegro | Montenegro | 83 | 0.278 | 0.261 |
| Morocco | Morocco | 75 | 0.244 | 0.246 |
| Mozambique | Mozambique | 105 | 0.095 | 0.151 |
| Nepal | Nepal | 108 | 0.109 | 0.201 |
| Netherlands | Netherlands | 64 | 0.486 | 0.471 |
| New Zealand | New Zealand | 79 | 0.342 | 0.346 |
| Niger | Niger | 90 | 0.373 | 0.427 |
| Nigeria | Nigeria | 82 | 0.184 | 0.178 |
| North Macedonia | North Macedonia | 77 | 0.297 | 0.298 |
| Norway | Norway | 62 | 0.334 | 0.332 |
| Oman | Oman | 78 | 0.137 | 0.163 |
| Pakistan | Pakistan | 72 | 0.474 | 0.539 |
| Panama | Panama | 73 | 0.251 | 0.300 |
| Paraguay | Paraguay | 84 | 0.182 | 0.180 |
| Peru | Peru_Amazonas | 103 | 0.161 | 0.236 |
| Peru | Peru_Ancash | 95 | 0.294 | 0.315 |
| Peru | Peru_Apurimac | 106 | 0.109 | 0.140 |
| Peru | Peru_Arequipa | 90 | 0.134 | 0.165 |
| Peru | Peru_Ayacucho | 106 | 0.153 | 0.208 |
| Peru | Peru_Cajamarca | 105 | 0.160 | 0.151 |
| Peru | Peru_Cusco | 89 | 0.108 | 0.097 |
| Peru | Peru_Huancavelica | 107 | 0.189 | 0.235 |
| Peru | Peru_Huanuco | 101 | 0.198 | 0.236 |
| Peru | Peru_Ica | 97 | 0.222 | 0.211 |
| Peru | Peru_Junin | 94 | 0.142 | 0.194 |
| Peru | Peru_La Libertad | 91 | 0.140 | 0.176 |
| Peru | Peru_Lambayeque | 90 | 0.319 | 0.363 |
| Peru | Peru_Lima | 73 | 0.283 | 0.283 |
| Peru | Peru_Loreto | 87 | 0.175 | 0.229 |
| Peru | Peru_Madre de Dios | 103 | 0.115 | 0.118 |
| Peru | Peru_Moquegua | 107 | 0.129 | 0.158 |
| Peru | Peru_Pasco | 109 | 0.161 | 0.183 |
| Peru | Peru_Piura | 91 | 0.243 | 0.309 |
| Peru | Peru_Puno | 111 | 0.200 | 0.187 |
| Peru | Peru_San Martin | 102 | 0.197 | 0.204 |
| Peru | Peru_Tacna | 108 | 0.129 | 0.121 |
| Peru | Peru_Tumbes | 94 | 0.159 | 0.171 |
| Peru | Peru_Ucayali | 104 | 0.186 | 0.177 |
| Philippines | Philippines | 70 | 0.243 | 0.253 |
| Poland | Poland | 71 | 0.336 | 0.343 |
| Portugal | Portugal | 68 | 0.348 | 0.423 |
| Qatar | Qatar | 71 | 0.102 | 0.163 |
| Romania | Romania | 70 | 0.398 | 0.387 |
| Russia | Russia_Adygea Republic | 105 | 0.284 | 0.334 |
| Russia | Russia_Altai Krai | 102 | 0.166 | 0.177 |
| Russia | Russia_Altai Republic | 118 | 0.048 | 0.079 |
| Russia | Russia_Amur Oblast | 114 | 0.111 | 0.150 |
| Russia | Russia_Arkhangelsk Oblast | 104 | 0.255 | 0.315 |
| Russia | Russia_Astrakhan Oblast | 103 | 0.298 | 0.270 |
| Russia | Russia_Bashkortostan Republic | 101 | 0.293 | 0.284 |
| Russia | Russia_Belgorod Oblast | 101 | 0.163 | 0.198 |
| Russia | Russia_Bryansk Oblast | 98 | 0.185 | 0.198 |
| Russia | Russia_Buryatia Republic | 92 | 0.083 | 0.078 |
| Russia | Russia_Chechen Republic | 102 | 0.313 | 0.281 |
| Russia | Russia_Chelyabinsk Oblast | 95 | 0.184 | 0.276 |
| Russia | Russia_Chuvashia Republic | 101 | 0.205 | 0.241 |
| Russia | Russia_Dagestan Republic | 93 | 0.172 | 0.239 |
| Russia | Russia_Ingushetia Republic | 99 | 0.431 | 0.496 |
| Russia | Russia_Irkutsk Oblast | 103 | 0.082 | 0.144 |
| Russia | Russia_Ivanovo Oblast | 97 | 0.137 | 0.144 |
| Russia | Russia_Jewish Autonomous Okrug | 112 | 0.173 | 0.182 |
| Russia | Russia_Kabardino-Balkarian Republic | 101 | 0.194 | 0.250 |
| Russia | Russia_Kaliningrad Oblast | 98 | 0.090 | 0.113 |
| Russia | Russia_Kalmykia Republic | 102 | 0.187 | 0.212 |
| Russia | Russia_Kaluga Oblast | 104 | 0.319 | 0.329 |
| Russia | Russia_Kamchatka Krai | 108 | 0.197 | 0.240 |
| Russia | Russia_Karachay-Cherkess Republic | 107 | 0.263 | 0.259 |
| Russia | Russia_Karelia Republic | 111 | 0.121 | 0.132 |
| Russia | Russia_Kemerovo Oblast | 109 | 0.192 | 0.183 |
| Russia | Russia_Khabarovsk Krai | 101 | 0.229 | 0.247 |
| Russia | Russia_Khakassia Republic | 106 | 0.259 | 0.265 |
| Russia | Russia_Khanty-Mansi Autonomous Okrug | 98 | 0.285 | 0.295 |
| Russia | Russia_Kirov Oblast | 100 | 0.175 | 0.206 |
| Russia | Russia_Komi Republic | 92 | 0.159 | 0.165 |
| Russia | Russia_Kostroma Oblast | 106 | 0.223 | 0.212 |
| Russia | Russia_Krasnodar Krai | 94 | 0.248 | 0.251 |
| Russia | Russia_Krasnoyarsk Krai | 93 | 0.202 | 0.303 |
| Russia | Russia_Kurgan Oblast | 116 | 0.087 | 0.093 |
| Russia | Russia_Kursk Oblast | 102 | 0.229 | 0.241 |
| Russia | Russia_Leningrad Oblast | 93 | 0.167 | 0.238 |
| Russia | Russia_Lipetsk Oblast | 97 | 0.187 | 0.211 |
| Russia | Russia_Magadan Oblast | 109 | 0.036 | 0.031 |
| Russia | Russia_Mari El Republic | 99 | 0.156 | 0.208 |
| Russia | Russia_Mordovia Republic | 103 | 0.236 | 0.324 |
| Russia | Russia_Moscow | 74 | 0.282 | 0.277 |
| Russia | Russia_Moscow Oblast | 85 | 0.266 | 0.296 |
| Russia | Russia_Murmansk Oblast | 99 | 0.246 | 0.253 |
| Russia | Russia_Nizhny Novgorod Oblast | 93 | 0.219 | 0.224 |
| Russia | Russia_North Ossetia - Alania Republic | 101 | 0.171 | 0.266 |
| Russia | Russia_Novgorod Oblast | 104 | 0.281 | 0.329 |
| Russia | Russia_Novosibirsk Oblast | 102 | 0.187 | 0.194 |
| Russia | Russia_Omsk Oblast | 108 | 0.105 | 0.137 |
| Russia | Russia_Orel Oblast | 102 | 0.226 | 0.263 |
| Russia | Russia_Orenburg Oblast | 101 | 0.285 | 0.299 |
| Russia | Russia_Penza Oblast | 94 | 0.164 | 0.195 |
| Russia | Russia_Perm Krai | 98 | 0.157 | 0.204 |
| Russia | Russia_Primorsky Krai | 104 | 0.193 | 0.215 |
| Russia | Russia_Pskov Oblast | 106 | 0.121 | 0.147 |
| Russia | Russia_Rostov Oblast | 101 | 0.304 | 0.315 |
| Russia | Russia_Ryazan Oblast | 99 | 0.258 | 0.264 |
| Russia | Russia_Saint Petersburg | 86 | 0.289 | 0.310 |
| Russia | Russia_Sakha (Yakutiya) Republic | 104 | 0.202 | 0.272 |
| Russia | Russia_Sakhalin Oblast | 124 | 0.059 | 0.106 |
| Russia | Russia_Samara Oblast | 104 | 0.150 | 0.189 |
| Russia | Russia_Saratov Oblast | 98 | 0.134 | 0.175 |
| Russia | Russia_Smolensk Oblast | 105 | 0.254 | 0.230 |
| Russia | Russia_Stavropol Krai | 98 | 0.162 | 0.166 |
| Russia | Russia_Sverdlovsk Oblast | 91 | 0.115 | 0.169 |
| Russia | Russia_Tambov Oblast | 101 | 0.232 | 0.233 |
| Russia | Russia_Tatarstan Republic | 94 | 0.154 | 0.194 |
| Russia | Russia_Tomsk Oblast | 110 | 0.137 | 0.142 |
| Russia | Russia_Tula Oblast | 96 | 0.144 | 0.223 |
| Russia | Russia_Tver Oblast | 102 | 0.317 | 0.340 |
| Russia | Russia_Tyumen Oblast | 102 | 0.260 | 0.260 |
| Russia | Russia_Tyva Republic | 117 | 0.161 | 0.250 |
| Russia | Russia_Udmurt Republic | 106 | 0.223 | 0.215 |
| Russia | Russia_Ulyanovsk Oblast | 101 | 0.191 | 0.186 |
| Russia | Russia_Vladimir Oblast | 100 | 0.248 | 0.255 |
| Russia | Russia_Volgograd Oblast | 98 | 0.140 | 0.188 |
| Russia | Russia_Vologda Oblast | 99 | 0.099 | 0.135 |
| Russia | Russia_Voronezh Oblast | 99 | 0.158 | 0.178 |
| Russia | Russia_Yamalo-Nenets Autonomous Okrug | 98 | 0.127 | 0.204 |
| Russia | Russia_Yaroslavl Oblast | 103 | 0.340 | 0.345 |
| Russia | Russia_Zabaykalsky Krai | 107 | 0.126 | 0.149 |
| Rwanda | Rwanda | 83 | 0.112 | 0.125 |
| Sao Tome and Principe | Sao Tome and Principe | 126 | 0.095 | 0.170 |
| Saudi Arabia | Saudi Arabia | 72 | 0.243 | 0.241 |
| Senegal | Senegal | 77 | 0.187 | 0.244 |
| Serbia | Serbia | 73 | 0.219 | 0.245 |
| Sierra Leone | Sierra Leone | 108 | 0.171 | 0.178 |
| Singapore | Singapore | 36 | 0.098 | 0.148 |
| Slovakia | Slovakia | 73 | 0.215 | 0.202 |
| Slovenia | Slovenia | 70 | 0.441 | 0.443 |
| South Africa | South Africa_Eastern Cape | 95 | 0.207 | 0.280 |
| South Africa | South Africa_Free State | 85 | 0.219 | 0.208 |
| South Africa | South Africa_Gauteng | 76 | 0.314 | 0.304 |
| South Africa | South Africa_Kwazulu-Natal | 81 | 0.313 | 0.333 |
| South Africa | South Africa_Limpopo | 106 | 0.060 | 0.138 |
| South Africa | South Africa_Mpumalanga | 118 | 0.112 | 0.120 |
| South Africa | South Africa_Northern Cape | 125 | 0.091 | 0.148 |
| South Africa | South Africa_North-West | 114 | 0.111 | 0.198 |
| South Africa | South Africa_Western Cape | 78 | 0.232 | 0.225 |
| South Sudan | South Sudan | 119 | 0.127 | 0.122 |
| Spain | Spain_Andalusia | 49 | 0.206 | 0.251 |
| Spain | Spain_Aragon | 60 | 0.227 | 0.223 |
| Spain | Spain_Asturias | 66 | 0.356 | 0.369 |
| Spain | Spain_Balearic Islands | 62 | 0.216 | 0.255 |
| Spain | Spain_Canary Islands | 59 | 0.204 | 0.214 |
| Spain | Spain_Cantabria | 65 | 0.237 | 0.256 |
| Spain | Spain_Castile and Leon | 54 | 0.231 | 0.239 |
| Spain | Spain_Castilla-La Mancha | 58 | 0.422 | 0.451 |
| Spain | Spain_Catalonia | 43 | 0.204 | 0.255 |
| Spain | Spain_Comunidad Valenciana | 46 | 0.195 | 0.252 |
| Spain | Spain_Extremadura | 59 | 0.214 | 0.270 |
| Spain | Spain_Galicia | 58 | 0.265 | 0.299 |
| Spain | Spain_La Rioja | 59 | 0.353 | 0.361 |
| Spain | Spain_Madrid | 34 | 0.165 | 0.247 |
| Spain | Spain_Murcia | 61 | 0.235 | 0.263 |
| Spain | Spain_Navarra | 63 | 0.330 | 0.355 |
| Spain | Spain_Pais Vasco | 56 | 0.321 | 0.329 |
| Sri Lanka | Sri Lanka | 76 | 0.240 | 0.223 |
| Sudan | Sudan | 104 | 0.302 | 0.311 |
| Sweden | Sweden_Blekinge | 92 | 0.038 | 0.087 |
| Sweden | Sweden_Dalarna | 83 | 0.222 | 0.230 |
| Sweden | Sweden_Gavleborg | 84 | 0.171 | 0.170 |
| Sweden | Sweden_Halland | 72 | 0.057 | 0.076 |
| Sweden | Sweden_Jamtland | 79 | 0.182 | 0.185 |
| Sweden | Sweden_Jonkoping | 72 | 0.093 | 0.129 |
| Sweden | Sweden_Kalmar lan | 87 | 0.086 | 0.104 |
| Sweden | Sweden_Kronoberg | 88 | 0.096 | 0.111 |
| Sweden | Sweden_Norrbotten | 84 | 0.136 | 0.129 |
| Sweden | Sweden_Orebro | 77 | 0.111 | 0.160 |
| Sweden | Sweden_Ostergotland | 75 | 0.272 | 0.300 |
| Sweden | Sweden_Skane | 70 | 0.486 | 0.467 |
| Sweden | Sweden_Sormland | 74 | 0.159 | 0.215 |
| Sweden | Sweden_Stockholm | 64 | 0.328 | 0.316 |
| Sweden | Sweden_Uppsala | 72 | 0.152 | 0.164 |
| Sweden | Sweden_Varmland | 72 | 0.048 | 0.078 |
| Sweden | Sweden_Vasterbotten | 80 | 0.107 | 0.146 |
| Sweden | Sweden_Vasternorrland | 87 | 0.197 | 0.211 |
| Sweden | Sweden_Vastmanland | 84 | 0.160 | 0.162 |
| Sweden | Sweden_Vastra Gotaland | 68 | 0.374 | 0.394 |
| Switzerland | Switzerland | 61 | 0.383 | 0.386 |
| Syria | Syria | 102 | 0.082 | 0.185 |
| Taiwan | Taiwan | 52 | 0.107 | 0.168 |
| Tajikistan | Tajikistan | 123 | 0.402 | 0.404 |
| Tanzania | Tanzania | 99 | 0.254 | 0.288 |
| Thailand | Thailand | 35 | 0.187 | 0.238 |
| Togo | Togo | 87 | 0.095 | 0.146 |
| Trinidad and Tobago | Trinidad and Tobago | 81 | 0.062 | 0.064 |
| Tunisia | Tunisia | 78 | 0.239 | 0.249 |
| Turkey | Turkey | 77 | 0.654 | 0.650 |
| Uganda | Uganda | 88 | 0.064 | 0.115 |
| Ukraine | Ukraine | 80 | 0.314 | 0.360 |
| United Arab Emirates | United Arab Emirates | 63 | 0.120 | 0.162 |
| United Kingdom | United Kingdom_East Midlands | 66 | 0.228 | 0.285 |
| United Kingdom | United Kingdom_East of England | 66 | 0.276 | 0.300 |
| United Kingdom | United Kingdom_London | 61 | 0.291 | 0.399 |
| United Kingdom | United Kingdom_North East | 74 | 0.226 | 0.248 |
| United Kingdom | United Kingdom_North West | 65 | 0.254 | 0.286 |
| United Kingdom | United Kingdom_Northern Ireland | 73 | 0.193 | 0.222 |
| United Kingdom | United Kingdom_Scotland | 70 | 0.375 | 0.386 |
| United Kingdom | United Kingdom_South East | 63 | 0.264 | 0.317 |
| United Kingdom | United Kingdom_South West | 64 | 0.183 | 0.239 |
| United Kingdom | United Kingdom_Wales | 71 | 0.358 | 0.347 |
| United Kingdom | United Kingdom_West Midlands | 67 | 0.351 | 0.370 |
| United Kingdom | United Kingdom_Yorkshire and The Humber | 67 | 0.237 | 0.245 |
| Uruguay | Uruguay | 76 | 0.333 | 0.330 |
| US | US_Alabama | 76 | 0.302 | 0.313 |
| US | US_Alaska | 83 | 0.204 | 0.220 |
| US | US_Arizona | 78 | 0.433 | 0.426 |
| US | US_Arkansas | 79 | 0.283 | 0.276 |
| US | US_California | 63 | 0.294 | 0.287 |
| US | US_Colorado | 71 | 0.347 | 0.352 |
| US | US_Connecticut | 76 | 0.389 | 0.396 |
| US | US_Delaware | 79 | 0.223 | 0.256 |
| US | US_Florida | 72 | 0.346 | 0.426 |
| US | US_Georgia | 72 | 0.321 | 0.316 |
| US | US_Hawaii | 79 | 0.215 | 0.213 |
| US | US_Idaho | 80 | 0.245 | 0.270 |
| US | US_Illinois | 71 | 0.363 | 0.466 |
| US | US_Indiana | 76 | 0.383 | 0.412 |
| US | US_Iowa | 78 | 0.207 | 0.228 |
| US | US_Kansas | 79 | 0.225 | 0.234 |
| US | US_Kentucky | 77 | 0.275 | 0.301 |
| US | US_Louisiana | 73 | 0.478 | 0.422 |
| US | US_Maine | 77 | 0.248 | 0.245 |
| US | US_Maryland | 75 | 0.344 | 0.346 |
| US | US_Massachusetts | 70 | 0.243 | 0.342 |
| US | US_Michigan | 74 | 0.439 | 0.462 |
| US | US_Minnesota | 76 | 0.212 | 0.222 |
| US | US_Mississippi | 78 | 0.452 | 0.464 |
| US | US_Missouri | 79 | 0.374 | 0.362 |
| US | US_Montana | 81 | 0.233 | 0.290 |
| US | US_Nebraska | 79 | 0.143 | 0.196 |
| US | US_Nevada | 76 | 0.255 | 0.290 |
| US | US_New Hampshire | 77 | 0.187 | 0.173 |
| US | US_New Jersey | 72 | 0.453 | 0.548 |
| US | US_New Mexico | 78 | 0.194 | 0.184 |
| US | US_New York | 66 | 0.403 | 0.544 |
| US | US_North Carolina | 74 | 0.355 | 0.371 |
| US | US_North Dakota | 80 | 0.228 | 0.233 |
| US | US_Ohio | 74 | 0.315 | 0.332 |
| US | US_Oklahoma | 79 | 0.301 | 0.361 |
| US | US_Oregon | 73 | 0.192 | 0.238 |
| US | US_Pennsylvania | 73 | 0.296 | 0.363 |
| US | US_Puerto Rico | 83 | 0.249 | 0.267 |
| US | US_Rhode Island | 78 | 0.203 | 0.197 |
| US | US_South Carolina | 76 | 0.352 | 0.344 |
| US | US_South Dakota | 83 | 0.183 | 0.200 |
| US | US_Tennessee | 73 | 0.322 | 0.371 |
| US | US_Texas | 72 | 0.323 | 0.404 |
| US | US_Utah | 76 | 0.314 | 0.338 |
| US | US_Vermont | 80 | 0.283 | 0.270 |
| US | US_Virginia | 73 | 0.223 | 0.243 |
| US | US_Washington | 63 | 0.262 | 0.405 |
| US | US_West Virginia | 85 | 0.266 | 0.274 |
| US | US_Wisconsin | 74 | 0.434 | 0.431 |
| US | US_Wyoming | 82 | 0.230 | 0.245 |
| Uzbekistan | Uzbekistan | 80 | 0.166 | 0.220 |
| Venezuela | Venezuela | 77 | 0.166 | 0.184 |
| Vietnam | Vietnam | 68 | 0.134 | 0.129 |
| West Bank and Gaza | West Bank and Gaza | 70 | 0.108 | 0.157 |
| Yemen | Yemen | 127 | 0.202 | 0.202 |
| Zambia | Zambia | 88 | 0.138 | 0.230 |
| Zimbabwe | Zimbabwe | 109 | 0.134 | 0.248 |

**Dataset S1.** Complete dataset, with data from all the regions included in the analyses (Excel file).

**Dataset S2.** Mean daily testing rate up to May 31, 2020 in 84 countries (COVID-19 tests performed per 1000 persons; Excel file).
